# SARS-CoV-2 recruits a haem metabolite to evade antibody immunity

**DOI:** 10.1101/2021.01.21.21249203

**Authors:** Annachiara Rosa, Valerie E. Pye, Carl Graham, Luke Muir, Jeffrey Seow, Kevin W. Ng, Nicola J. Cook, Chloe Rees-Spear, Eleanor Parker, Mariana Silva dos Santos, Carolina Rosadas, Alberto Susana, Hefin Rhys, Andrea Nans, Laura Masino, Chloe Roustan, Evangelos Christodoulou, Rachel Ulferts, Antoni Wrobel, Charlotte-Eve Short, Michael Fertleman, Rogier W. Sanders, Judith Heaney, Moira Spyer, Svend Kjær, Andy Riddell, Michael H. Malim, Rupert Beale, James I. MacRae, Graham P. Taylor, Eleni Nastouli, Marit J. van Gils, Peter B. Rosenthal, Massimo Pizzato, Myra O. McClure, Richard S. Tedder, George Kassiotis, Laura E. McCoy, Katie J. Doores, Peter Cherepanov

**Author notes:** Corresponding authors. (PC); (KJD); (LEM); (GK).

## Abstract

The coronaviral spike is the dominant viral antigen and the target of neutralizing antibodies. We show that SARS-CoV-2 spike binds biliverdin and bilirubin, the tetrapyrrole products of haem metabolism, with nanomolar affinity. Using cryo-electron microscopy and X-ray crystallography we mapped the tetrapyrrole interaction pocket to a deep cleft on the spike N-terminal domain (NTD). At physiological concentrations, biliverdin significantly dampened the reactivity of SARS-CoV-2 spike with immune sera and inhibited a subset of neutralizing antibodies. Access to the tetrapyrrole-sensitive epitope is gated by a flexible loop on the distal face of the NTD. Accompanied by profound conformational changes in the NTD, antibody binding requires relocation of the gating loop, which folds into the cleft vacated by the metabolite. Our results indicate that the virus co-opts the haem metabolite for the evasion of humoral immunity via allosteric shielding of a sensitive epitope and demonstrate the remarkable structural plasticity of the NTD.

---

Trimeric coronaviral spike glycoproteins form prominent features on viral particles that are responsible for the attachment to a receptor on the host cell and, ultimately, fusion of the viral and cellular membranes (*1, 2*). Encoded by a single viral gene, the mature spike glycoprotein comprises two subunits, S1 and S2, which mediate binding to the receptor and facilitate fusion, respectively. The recognition of the betacoronavirus SARS-CoV-2 host receptor, the cellular membrane protein angiotensin-converting enzyme 2, maps to the S1 C-terminal domain (referred to as the receptor binding domain, RBD) (*3-5*), while the function of the N-terminal domain (NTD) remains enigmatic. Both domains can be targeted by potent neutralizing antibodies that arise in infected individuals. The majority of characterized neutralizing antibodies bind the RBD, while minimal structural information exists about neutralizing epitopes on the NTD (*6-10*). The immune properties of the spike glycoprotein underpin ongoing SARS-CoV-2 vaccine development efforts (*11*).

In the course of our activities to support the development of serology for SARS-CoV-2, we produced a range of recombinant coronaviral spike antigens by expression in human cell lines (Fig. S1A). Surprisingly, preparations of SARS-CoV-2 trimeric spike and S1 carried a distinctive green hue, with prominent peaks at ∼390 and 670 nm in their light absorbance spectra (Fig. S1A-B). These unusual features were also evident in the spectrum of S1 from the 2003 SARS-CoV-1 isolate, but not those from the seasonal human coronaviruses NL63 and OC43 (Fig. S1B-C). The property was confined within the spike NTD and absent in isolated RBD (Fig. S1B). The spectra of the SARS-CoV spike constructs were consistent with biliverdin (Fig. S1B), a product of haem metabolism responsible for the coloration of bruises and green jaundice. We isolated the pigment from denatured SARS-CoV-2 S1 and confirmed the presence of biliverdin IXα by mass spectrometry (Fig. S2). Biliverdin is produced at the first step of haem detoxification by oxygenases and is then reduced to bilirubin, the final product of tetrapyrrole catabolism in humans (*12*). We measured tetrapyrrole binding to immobilised SARS-CoV-2 S1 using surface plasmon resonance (SPR) and estimated the dissociation constant (K_d_) for the interaction with biliverdin and bilirubin at 9.8 ±1.3 nM and 720 ±250 nM, respectively (Fig. S3A-B, Table S1). SARS-CoV-1 S1 likewise displayed nanomolar affinity for biliverdin (Fig. S3I, Table S1). The spike bound haem considerably more weakly, with K_d_ of 7.0 ±1.2 μM, while no interaction was observed with protoporphyrin IX (Fig. S3C-D, Table S1).

Next, we imaged single particles of the trimeric SARS-CoV-2 spike ectodomain (*3, 13*) in the presence of excess biliverdin using cryo-electron microscopy. Image processing resulted in the 3D reconstruction of closed (3RBDs-down) and partially open (1RBD-up conformation) states of the spike at 3.35 and 3.50 Å resolution, respectively (Fig. 1A, Fig. S4, Table S2). Close inspection of the cryo-EM maps revealed features interpretable as a biliverdin molecule buried within a deep cleft on one side of each of the NTD domains (Fig. 1A, Fig. S5A). To define the structural basis for the interaction more precisely, we co-crystallised the isolated NTD with biliverdin and determined the structure at 1.8 Å resolution (Fig. 1B, Fig. S5B, Table S3). The metabolite fits snugly into the cleft with the pyrrole rings B and C buried inside and propionate groups appended to rings A and D projecting toward the outside. The pocket is lined by hydrophobic residues (Ile101, Trp104, Ile119, Val126, Met177, Phe192, Phe194, Ile203, and Leu226), which form van der Waals interactions with the ligand. Biliverdin packs against His207, which projects its Nε2 atom towards pyrrolic amines, approaching three of them at ∼3.6 Å. Pyrroles A and B are involved in a π-π stacking with side chain of Arg190, which is stabilised by hydrogen bonding with Asn99. The binding of biliverdin largely buries the side chain of Asn121, which makes a hydrogen bond with the lactam group of pyrrole D. In agreement with the extensive interactions observed in the crystal structure, the melting point of isolated NTD increased by over 8°C in the presence of biliverdin (Fig. S3K). Unidentified entities at the tetrapyrrole binding site were observed in published SARS-CoV-2 spike reconstructions (*1, 4, 13-17*), presumably obtained with partial occupancy by the metabolite; in some cryo-EM maps, the bound biliverdin molecule is resolved remarkably well (Fig. S6) (*14-17*).

**Figure 1.**
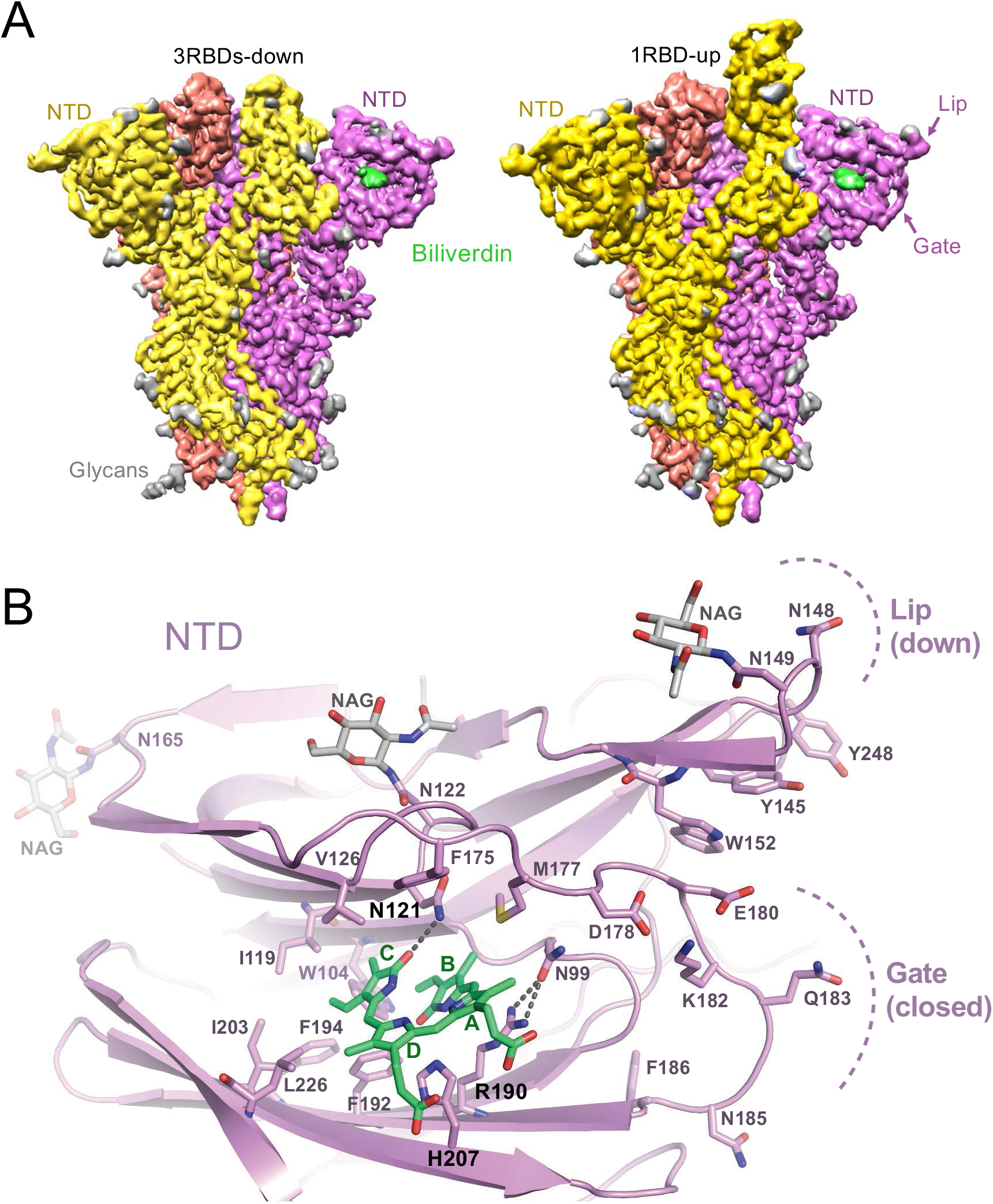
Structures of SARS-CoV-2 spike-biliverdin (A,B) and spike-P008_056 Fab (C) complexes. (**A**) Cryo-EM 3D reconstructions of trimeric SARS-CoV-2 spike ectodomain in 3RBD-down (left) and 1RBD-up (right) conformations determined under saturation with biliverdin. Spike protomers are color-coded. Biliverdin and glycans are shown in green and grey, respectively. (**B**) Details of the biliverdin binding pocket in the crystal structure. SARS-CoV-2 NTD is shown as cartoons with selected amino acid residues and biliverdin in sticks. Carbon atoms of the protein chain, sugars (NAG), and biliverdin are in purple, grey and green, respectively; the remaining atoms are coloured as follows: oxygen, red; nitrogen, blue; and sulphur, yellow. Dark grey dashes are hydrogen bonds.

The presence of a histidine residue in the biliverdin binding pocket (Fig. 1B) suggested that the interaction may be pH-dependent. In agreement with this hypothesis, the K_d_ of the S1-biliverdin interaction increased to 250 ±100 μM at pH 5.0 (Fig. S3E,H and Table S1), and purification under acidic conditions greatly reduced the biliverdin content of recombinant SARS-CoV-2 S1 (Fig. S1D). Substitutions of spike residues closely involved in ligand binding (H207A, R190K and N121Q) diminished pigmentation of purified recombinant protein (Fig. S1E). The biliverdin binding affinity of SARS-CoV-2 S1 was reduced by two and three orders of magnitude by the R190K and N121Q amino acid substitutions, respectively (Fig. S3F-H, Table S1). The latter ablated the interaction with bilirubin, confirming that the tetrapyrroles share the binding site on the spike (Fig. S3H). SARS-CoV-2 spikes carrying these mutations supported infection of Vero and Huh7 cells by a pseudotyped retroviral vector, indicating that biliverdin binding is not required for viral entry under tissue culture conditions (Fig. S7).

Because biliverdin binding conceals a deep hydrophobic cleft on the NTD (Fig. 1B), we suspected that it may mask or modify the antigenic properties of the viral spike. To test this hypothesis, we measured the reactivity of sera from 17 SARS-CoV-2-infected and convalescent individuals with full-length WT and N121Q SARS-CoV-2 spike using a flow cytometry-based assay (*18*). Remarkably, addition of 10 μM biliverdin reduced binding of patient IgGs to WT spike, supressing the reactivity of some of the immune sera by as much as 50% (Fig. 2). By contrast, antibody binding to N121Q SARS-CoV-2 spike was not affected by addition of biliverdin (Fig. 2). Binding of IgM and IgA antibodies, which are present at lower titres in these patients (*18*), was more variably affected (Fig. S8B-D). In a separate experiment, we tested 91 clinical serum samples in an IgG capture enzyme-linked immunosorbent assay (ELISA). SARS-CoV-2 specific antibodies were detected using WT or N121Q S1 conjugated to horseradish peroxidase. Biliverdin-depleted WT S1 was significantly more reactive than the same protein supplemented with excess metabolite (Fig. S9). By contrast, addition of biliverdin did not dampen detection of the SARS-CoV-2 antibodies with N121Q S1 (Fig. S9).

**Figure 2.**
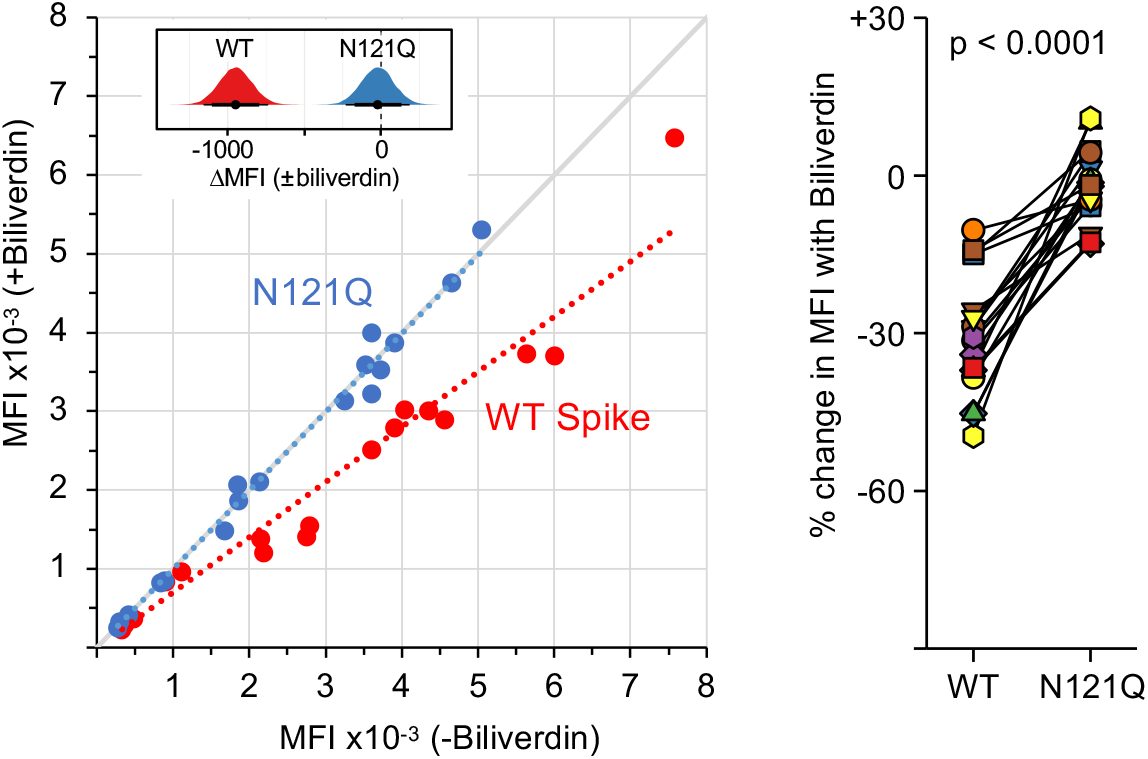
Biliverdin strongly downmodulates the reactivity of SARS-CoV-2 spike with antibodies present in immune sera. *Left:* Mean fluorescence intensity (MFI) of IgG staining of HEK293T cells expressing full-length WT or N121Q SARS-CoV-2 spike by individual patient sera in the absence or the presence of 10 µM biliverdin. Each symbol represents an individual patient (n=17) and coloured dotted lines represent the linear regression for each spike variant. The inset shows posterior probability density plots of values for pairwise contrasts (±biliverdin) for the WT and N121Q spikes. Black dots indicate the median of the distribution, thick and thin line ranges correspond to the 85% and 95% highest density interval, respectively; the dotted vertical line indicates a zero difference. *Right:* Changes in MFI caused by the addition of 10 μM biliverdin, as percent of staining without biliverdin, for serum for IgM and IgA antibodies. Each pair of connected symbols represents an individual patient.

It is remarkable that a small molecule with a footprint of 370 Å^2^, corresponding to only ∼0.9% of solvent-exposed surface (per spike monomer, Fig. 1A), competes with a considerable fraction of the spike-specific serum antibody population (Fig. 2). These results prompted us to evaluate a panel of human antibodies cloned from B cells of SARS-CoV-2 convalescent individuals. We used 38 IgGs reported in a recent study (*10*), as well as a panel of 15 novel SARS-CoV-2 S1-specific IgGs obtained from individuals with asymptomatic infection, or mild/severe disease undergoing characterization in one of our laboratories (Graham *et al*., in preparation). We tested these monoclonal antibodies for binding to recombinant WT and N121Q S1 by ELISA. Remarkably, 9 of 53 (17%) IgGs lost binding to WT, but not to N121Q S1, in the presence of 10 μM biliverdin (Fig. 3A,C, Fig. S10A). Furthermore, addition of biliverdin strongly suppressed binding of these antibodies to full-length WT but not N121Q SARS-CoV-2 spike expressed on the surface of transfected HEK293T cells (Fig. 3B,D). By contrast, reactivity of both RBD-specific controls (COVA1-18 and COVA1-12), was not affected by the metabolite (Fig. 3A-D).

**Figure 3.**
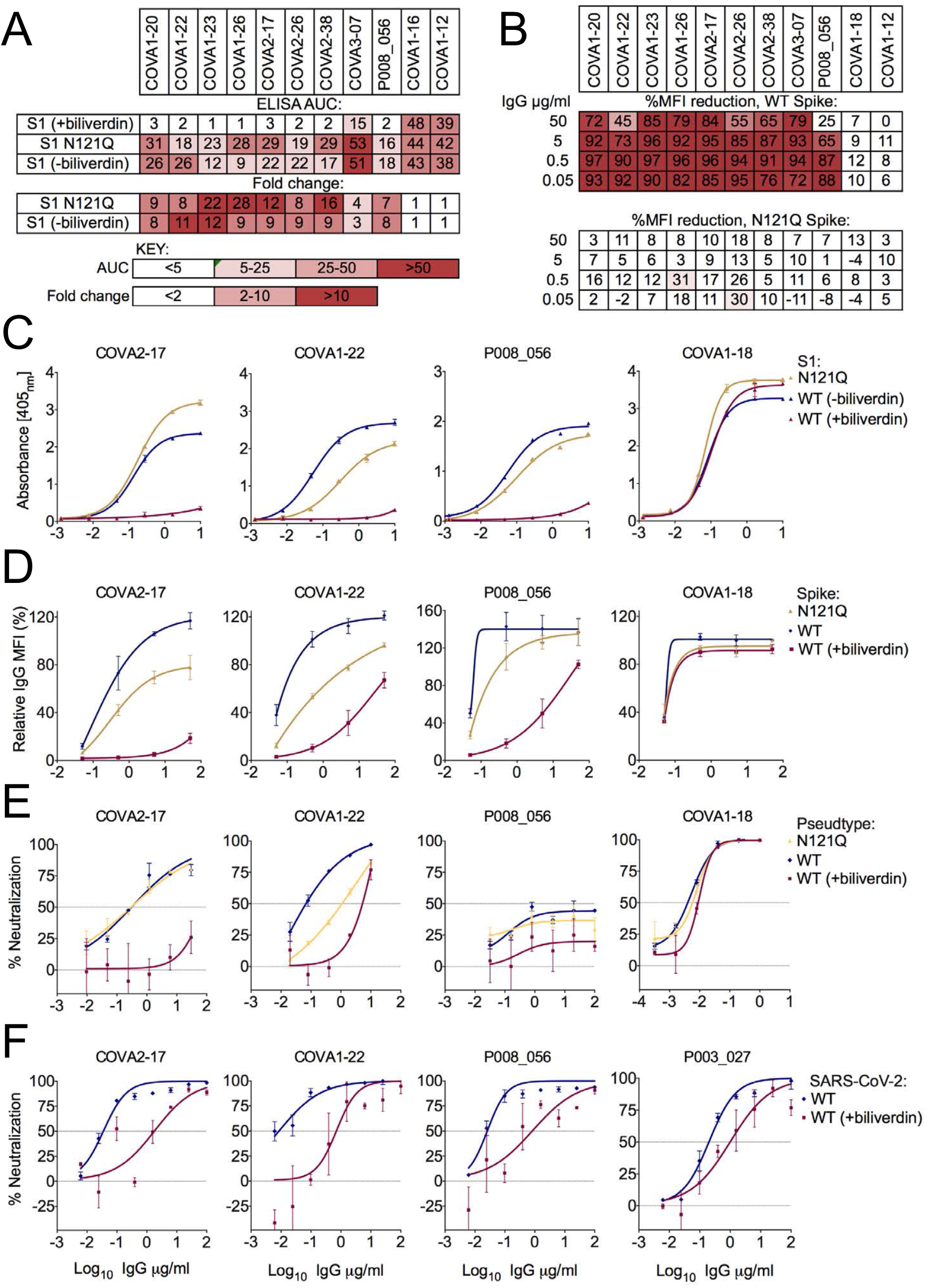
Biliverdin decreases binding to SARS-CoV-2 spike by a group of human monoclonal IgGs. (**A**) Antibodies were titrated 6-fold and assayed by direct ELISA for binding to recombinant S1 biliverdin-depleted by purification under acidic conditions (-biliverdin), same protein but supplemented with biliverdin (+biliverdin) or N121Q S1. Area under the curve (AUC) is shown for IgG that were sensitive to biliverdin and two unaffected control IgGs. AUC values are colour-coded as per the key; fold change compared to WT protein are reported. (**B**) Biliverdin-sensitive IgGs were titrated 10-fold and incubated with 293T cells expressing full-length WT or N121Q SARS-CoV-2 spike with or without 10 μM biliverdin. Binding was detected using an anti-IgG antibody and reduction in binding in the presence of biliverdin is shown as % MFI reduction and colour-coded as a heatmap of the quartile values. (**C**) ELISA titration curves for four neutralizing IgG including the biliverdin-insensitive control COVA1-18. (**D**) Relative MFI dose-dependent curves for four neutralizing IgG including the biliverdin insensitive control COVA1-18. Relative MFI calculated by normalising to the MFI of the biliverdin-insensitive COVA1-18 at the highest concentration against spike. (**E**) IgG indicated above each graph were titrated 5-fold against SARS-CoV-2 spike pseudotype, in the presence and absence of 10 μM biliverdin, and a version of spike encoding the mutation N121Q. COVA1-18 was used as a biliverdin-insensitive control IgG. (**F**) Neutralization of SARS-CoV-2 (England 02/2020/407073) by IgGs was measured in the absence and presence of 10 μM biliverdin in Vero-E6 cells. P003_027 was used as a biliverdin-insensitive control IgG.

Two of the biliverdin-sensitive monoclonal IgGs, COVA1-22, COVA2-17, were previously reported to efficiently neutralize SARS-CoV-2 pseudotyped retrovirus (*10*). Addition of biliverdin suppressed neutralization of the pseudotype carrying WT but not N121Q spike (Fig. 3E). The mutation had a differential effect on neutralization showing a decrease in potency for COVA1-22 and no effect for P008_056 or COVA2-17. As expected, addition of biliverdin had no effect on neutralization by COVA1-18. Neutralization of replication-competent SARS-CoV-2 by P008_056, COVA1-22 or COVA2-17 was substantially decreased by addition of biliverdin (Fig. 3F). Intriguingly, while P008_056 seemed somewhat less potent in the pseudotype assay (Fig. 3E), it efficiently neutralized live SARS-CoV-2 virus, achieving 50% and >90% inhibition at concentrations of 0.03 and 1.56 μg/ml, respectively, in a biliverdin-sensitive manner (Fig. 3F, Fig. S10B).

To establish the structural basis for SARS-CoV-2 neutralization by a biliverdin-sensitive antibody, we imaged single particles of the viral spike in complex with P008_056 antigen binding fragment (Fab). Cryo-EM image processing resulted in reconstruction of structures with one, two and three Fab moieties bound per trimeric viral glycoprotein (Fig. S10A-B), and the best map was obtained for the complex containing a single Fab (Fig. 4A, Fig. S11C-D). The reconstruction with a local resolution of ∼4 Å at the NTD-Fab interface allowed for unambiguous tracing of the protein backbone and revealed positions of key amino acid side chains (Fig. 4A, Fig. S5C). P008_056 binds the spike at the side of the NTD β-sandwich fold, which undergoes profound conformational rearrangements (Movie S1). Access to the epitope is gated by a solvent-exposed loop composed of predominantly hydrophilic residues (“gate”, SARS-CoV-2 spike residues 174-188; Fig. 1). To allow P008_056 binding, the loop swings out of the way, with a backbone displacement in the middle of the loop of ∼15 Å (Fig. 4B). The gating mechanism is accompanied by insertion of Phe175 and Met177, which are located in the beginning of the loop, into the hydrophobic pocket vacated by biliverdin (Fig. 4B, Fig. S5C). Thus, when bound, the metabolite appears to act as a wedge that restricts gate opening. Antibody binding is additionally complemented by an upward movement of a β-hairpin (“lip”, SARS-CoV-2 residues 143-155), which overlays a cluster of aromatic residues (Fig. 1, Fig. 4B). The marked gain in thermal stability upon biliverdin binding (Fig. S3K) is consistent with resistance to an antibody that requires major conformational remodeling of the NTD for binding (Movie S1).

**Figure 4.**
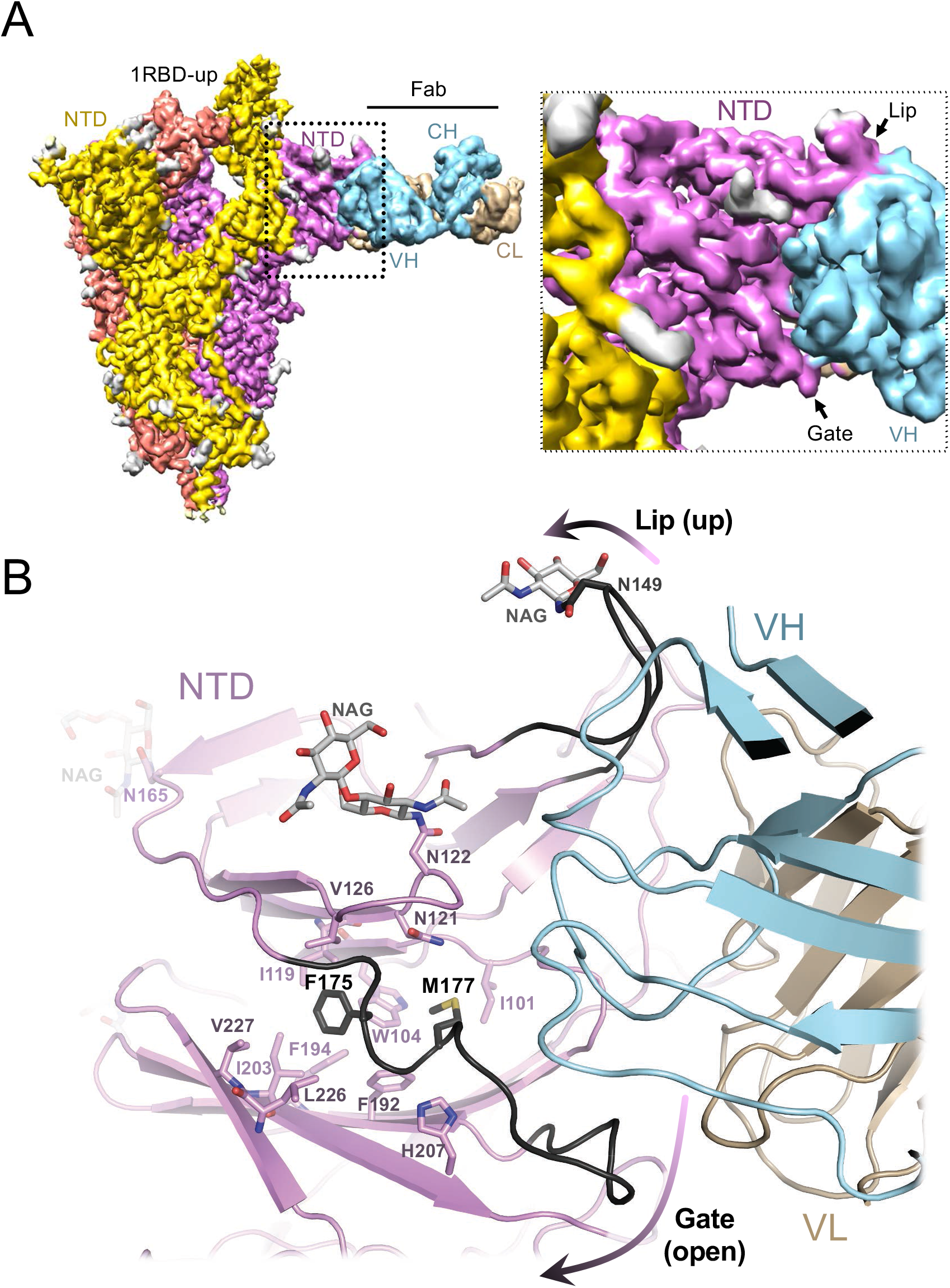
Cryo-EM structure of the spike-Fab complex. (**A**) Reconstruction obtained with multibody refinement in Relion (*left*) and a zoom on the spike-Fab interface in the structure obtained by consensus refinement (Fig. S11d). (**B**) Refined model of the spike-Fab complex shown as cartoon, with selected amino acid side chains in sticks and indicated. Carbon atoms of the gate and lip NTD elements that relocate to allow Fab binding (arrows), are shown in black. Fab heavy (HV) and light (LV) chains are shown in blue and beige, respectively.

It is well-established that viruses employ extensive glycosylation of their envelopes to shield antibody epitopes from recognition by humoral immunity (*19-21*). Here, we identified and structurally characterised a novel class of a neutralizing epitope, present on SARS-CoV-2 spike, which is differentially exposed through recruitment of a metabolite. In contrast to glycosylation, co-opting a metabolite may allow conditional unmasking, for example under acidic conditions within the endosomal compartment. Biliverdin levels in plasma of healthy individuals (0.9-6.9 μM) and more so under pathological conditions (>50 μM) (*22*) greatly exceed the K_d_ of its interaction with the spike (∼10 nM) and are therefore sufficient to affect SARS-CoV-2 antigenic properties and neutralization. Although SARS-CoV-2 spike bound bilirubin with lower affinity (Fig. S3), this final product of haem catabolism accumulates at higher levels *in vivo* (*22*). Moreover, elevated bilirubin levels correlate with the symptoms and mortality among COVID-19 patients (*23-26*). Therefore, the tetrapyrroles likely share a role in SARS-CoV-2 immune evasion. Severe COVID-19 symptoms and death are associated with neutrophil infiltration in pulmonary capillaries and alveolar space (*27*). Indeed, nasopharyngeal swabs of COVID-19 patients are enriched in neutrophil myeloperoxidase (*28*), a highly abundant haem-containing protein responsible for coloration of mucus (*29*). Alongside extensive vascular damage, these symptoms provide rich source of haem catabolites, which may contribute to the inability to control the infection in severe cases. Mutations within the SARS-CoV-2 spike NTD are associated with viral escape from antibody immunity (*30, 31*) and have been observed in circulating viral strains (*32, 33*). Our results demonstrate a remarkable structural plasticity of the NTD and highlight the importance of this domain for antibody immunity against SARS-CoV-2.

## Data Availability

The crystal structure of the NTD in complex with biliverdin and associated X-ray diffraction data are deposited with the Protein Data Bank under accession code 7B62. The cryo-EM maps and refined models of the trimeric SARS-CoV-2 spike will be released prior to publication of the peer-reviewed manuscript. Further information and requests for unique reagents should be directed to the corresponding authors.

## Acknowledgements

We are grateful to A.N. Engelman, A. Hayday, J.P. Stoye, and J. Skehel for comments on the manuscript, D. Benton and G.N. Maertens for helpful discussions, P. Walker and A. Purkiss for computer, software, and synchrotron access support, the staff of the Swiss Light Source PX1 beamline for assistance with data collection, Chris Wilson and Atul Goyale (UCLH Biochemistry) for their assistance with serum sample separation, and J. E. Voss for the kind gift of HeLa cells stably expressing ACE2. This research was funded by the Francis Crick Institute (PC), which receives its core funding from Cancer Research UK, the UK Medical Research Council, and the Wellcome Trust. It was funded also by the US National Institutes of Health (AI150481, PC) the King’s Together Rapid COVID-19 Call award (KJD and MHM), the Huo Family Foundation (MHM and KJD) and by the UCL Coronavirus Response Fund made possible through generous donations from UCL supporters, alumni and friends (LEM, LM). Flow cytometry was supported by a multi-user equipment grant from The Wellcome Trust to MHM and KJD (208354/Z/17/Z). LEM is supported by a Medical Research Council Career Development Award (MR/R008698/1). MJvG is a recipient of an AMC Fellowship, and RWS is a recipient of a Vici grant from the Netherlands Organization for Scientific Research (NWO). CG was supported by the MRC-KCL Doctoral Training Partnership in Biomedical Sciences (MR/N013700/1). MOM, EP and RST were supported in part by an UKRI/MRC Covid-19 grant (MC_PC_19078). EN, MJS, JH receive support from the UKRI COVID-19 research scheme. This study is part of the EDCTP2 program supported by the European Union (grant number RIA2020EF-3008 COVAB) (KJD and MHM). The views and opinions of authors expressed herein do not necessarily state or reflect those of EDCTP. This research was funded in whole, or in part, by the Wellcome Trust. For the purpose of Open Access, the author has applied a CC BY public copyright license to any author accepted manuscript version arising from this submission.

## Author contributions

AR, CR, EC, AW, and PC expressed and purified proteins; CR and SK established a stable cell line secreting trimeric SARS-CoV-2 spike; MSdS and JIMR conducted mass spectrometry; AR and VEP crystallised NTD-biliverdin complex; AN and PBR acquired and evaluated cryo-EM data; VEP collected and processed X-ray diffraction data, built and refined all atomistic models; AR and PC refined cryo-EM structures; NC and LM conducted SPR measurements; AR performed thermostability assays; ELP, CRdO, MOM, and RST performed IgG capture ELISA; HR and AR did statistical modelling of IgG capture assay data; RU and RB evaluated SARS-CoV2 infectivity in the presence of biliverdin; AS and MP conducted pseudotype infectivity assays; C-ES, MF, JH, MS, GPT, and EN assembled the panels of human sera samples; MJvG, RWS, KJD, JS, and CG isolated or provided monoclonal antibodies; KWN and GK did flow cytometry assays with human sera; KJD, JS, CG, ELP, LEM, LMuir, MHM, and CR-S characterised monoclonal antibodies; PC, KJD, LEM, and GK wrote the paper with contributions from all authors.

## Competing interests

The authors declare no competing interests.

## Materials and Methods

### Protein expression and purification

DNA fragments encoding SARS-CoV-2 S1 (Uniprot ID: P0DTC2; residues 1-530), NTD (1-310), RBD (319-541), SARS-CoV-1 S1 (Uniprot ID: P59594; residues 1-518), HcoV NL63 (Uniprot ID: Q6Q1S2; residues 1-618), HCoV OC43 (isolate LRTI_238, NCBI accession code KX344031; residues 1-619) were codon-optimised for expression in human cells and cloned into pQ-3C2xStrep vector (*34*) under control of the cytomegalovirus (CMV) promoter for production of the recombinant proteins carrying a C-terminal extension containing human rhinovirus 14 3C protease recognition site followed by a TwinStrep tag. The signal peptide from immunoglobulin kappa gene product (METDTLLLWVLLLWVPGSTGD) was used to direct secretion of the RBD construct. The vector for production of the His_6_-tagged stabilised trimeric SARS-CoV-2 has been described (*13*). Expression constructs encoding heavy and light chains of P008_056 Fab were made by inserting the respective coding sequences into pHLsec (*35*), including a sequence encoding a hexa-histidine (His_6_) tag on the heavy chain fragment C-terminus.

With exception of trimeric stabilized SARS-CoV-2 spike ectodomain, the proteins were produced by transient transfection of Expi293 (Thermo Fisher Scientific) cells with endotoxin-free preparations of the corresponding DNA constructs using ExpiFectamine293 (Thermo Fisher Scientific). The cells were maintained in shake flasks in FreeStyle293 (Thermo Fisher Scientific) medium at 37°C in humidified 5% CO_2_ atmosphere. To produce SARS-CoV-2 S1 NTD fragment for crystallography, cell culture medium was supplemented with 5 μM kifunensine (Sigma-Aldrich) to suppress complex glycosylation (*36, 37*). Conditioned medium containing recombinant product was harvested twice, 4 and 8 days post-transfection, or once, for production of the NTD and P008_056 Fab, 5 days post-transfection. For production of the trimeric SARS-CoV-2 spike ectodomain, Expi293 transfected with the pcDNA3-based expression construct (*13*) were selected with 250 μg/ml geneticin. Stably transfected cells, grown to a density of 3.5 million per ml at 37°C, were shifted to 32°C for 3 days prior to harvesting conditioned medium to enhance secretion of the viral glycoprotein (*38*).

TwinStrep-tagged proteins were captured on Strep-Tactin XT (IBA LifeSciences) affinity resin. Following extensive washes in TBSE (150 mM NaCl, 1 mM ethylenediaminetetraacetic acid (EDTA), 25 mM Tris-HCl, pH 8.0), the proteins were eluted in 1xBXT buffer (IBA LifeSciences). His_6_-tagged proteins were captured on HisTrap Excel (Sigma-Aldrich) resin and eluted with 300 mM imidazole in phosphate buffered saline. For the use in crystallography, SARS-CoV-2 S1 NTD was digested with Endo Hf (New England Biolabs) and rhinoviral 3C protease to trim glycans and to remove the C-terminal twin Strep tag; Endo Hf was depleted by absorption to amylose resin (New England Biolabs). The proteins were further purified by size exclusion chromatography through a Superdex 200 16/600 column (GE Healthcare) in HBSE (150 mM NaCl, 1 mM EDTA, 20 mM Hepes-NaOH, pH 8.0) and concentrated by ultrafiltration using a Vivaspin-20 with 10-kDa cut-off (Sartorius). To deplete biliverdin from SARS-CoV-2 S1, recombinant protein eluted from Strep-Tactin XT resin was supplemented with 0.5 M sodium acetate, pH 5.2 and subjected to size exclusion chromatography through a Superdex 200 16/600 column in 200 mM sodium acetate, pH 5.2; fractions containing S1 were pooled and dialyzed overnight against HBSE buffer. Light absorbance spectra of recombinant proteins were acquired using Jasco V-550 UV/VIS spectrophotometer.

### Identification of biliverdin by mass spectrometry

Recombinant SARS-CoV-2 S1 (3.5 mg/ml, 500 μl), denatured by addition of 1% (w/v) sodium dodecyl sulfate, was extracted with 500 μl n-butanol. Organic phase containing green pigment was allowed to evaporate under vacuum, and dry residue was re-dissolved in 20 μl water. Liquid chromatography-tandem mass spectrometry was performed as described previously (*39*). LC-MS/MS analysis was conducted using a Dionex UltiMate LC system (Thermo Scientific) with a ZIC-pHILIC column (150 mm x 4.6 mm, 5 μm particle, Merck Sequant). A 15-min elution gradient of 80% Solvent A (20 mM ammonium carbonate in Optima HPLC grade water, Sigma-Aldrich) to 20% Solvent B (acetonitrile Optima HPLC grade, Sigma-Aldrich) was used, followed by a 5-min wash of 95:5 Solvent A to Solvent B and 5-min re-equilibration. Other parameters were as follows: flow rate, 300 μl/min; column temperature, 25°C; injection volume, 10 μl; autosampler temperature, 4°C. Metabolites were detected across a mass range of 70-1050 m/z using a Q Exactive Orbitrap instrument (Thermo Scientific) with heated electrospray ionization and polarity switching mode at a resolution of 70,000 (at 200 m/z). MS parameters were as follows: spray voltage 3.5 kV for positive mode and 3.2 kV for negative mode; probe temperature, 320°C; sheath gas, 30 arbitrary units; auxiliary gas, 5 arbitrary units. Parallel reaction monitoring was used at a resolution of 17,500 to confirm the identification of biliverdin; collision energy was set at 30 in high-energy collisional dissociation mode. Data was recorded using Xcalibur 3.0.63 software and analysed using Free Style 1.6 and Tracefinder 4.1 software (Thermo Scientific) according to the manufacturer’s workflows.

### SPR

Experiments were performed on a Biacore S200 (Cytiva); S1 protein, diluted to 50 μg/ml in 10 mM sodium acetate, pH 5.0, was immobilized on a CM5 sensor chip (Cytiva product code BR100530) using amine coupling chemistry. Immobilization levels were typically 4,000 response units. Analyte binding was studied in running buffer comprising 150 mM NaCl, 50 mM HEPES-NaOH, pH 8.0 or 50 mM BisTris-HCl, pH 5.0, 0.05% Tween-20, and 1% dimethyl sulfoxide (DMSO). Biliverdin, bilirubin, haem, and protoporphyrin were obtained from Sigma-Aldrich (product codes 3089, 14370, 51280, and P8293, respectively). Generally, analyte stock solutions were prepared in DMSO prior to dilution in running buffer, maintaining the final DMSO concentration of 1%. The final analyte concentration was verified by spectrophotometry, using the following molar extinction coefficients: biliverdin 39,900 (at a wavelength of 388 nm), bilirubin 53,846 (460 nm), haem 58,440 (385 nm), and protoporphyrin IX 107,000 (407 nm). Alternatively, biliverdin, which is highly soluble at pH>7, was dissolved directly in running buffer, allowing to omit DMSO from the experiment. The presence of DMSO did not affect the measured K_d_ of the S1-biliverdin interaction (Table S1). All experiments were conducted using a CM5-kinetics-multicycle template at 25°C. Flow rate was 30 μl/min with a contact time of 180 s, followed by a dissociation time of 10 min; three start-ups were performed at the beginning of each experiment. Solvent correction was deemed unnecessary for the assays that contained DMSO. Biliverdin displayed very fast association precluding detailed kinetics analyses due to mass transfer effects. Data were analysed using the 1:1 binding model in the Biacore S200 Evaluation software to estimate equilibrium binding constants (K_d_).

### Protein thermostability assay

Biliverdin-depleted SARS-CoV-2 NTD (corresponding to spike residues 1-310) was diluted to 1 mg/ml in 150 mM NaCl, 20 mM HEPES-NaOH, pH 8.0 and supplemented with biliverdin from a 5-mM stock prepared in 100 mM Tris-HCl, pH8.0 where appropriate. Melting curves were recorded using 20−95 °C 1.5 °C/min temperature ramps on a Promethius NT.48 instrument (Nanotemper). Melting points were determined from inflection points of fluorescence intensity ratios (350 and 330 nm) using first derivative analysis (fig. S3K).

#### Cryo-electron microscopy

Four μl stabilised trimeric SARS-CoV-2 spike ectodomain (0.6 mg/ml final concentration in TBSE supplemented with 0.1% n-octylglucoside) with 25 μM biliverdin or 0.2 mg/ml P008_056 Fab, was applied onto glow-discharged 200-mesh copper holey carbon R2/2 grids (Quantifoil) for 1 min, under 100% humidity at 20°C, before blotting for 3-4 sec and plunge-freezing in liquid ethane using Vitrobot Mark IV (Thermo Fisher Scientific). The data were collected on Titan Krios microscopes operating at 300 keV (Thermo Fisher Scientific). Single particles of spike-biliverdin were imaged using a Falcon III direct electron detector (Thermo Fisher Scientific). A total of 15,962 movies were recorded with a calibrated pixel size of 1.09 Å and a total electron exposure of 33 e^-^/Å^2^, spread over 30 frames in single electron counting mode. The spike-Fab complex was imaged on a GIF Quantum K2 detector with a post-column energy filter (Gatan), selecting a 20-eV window, in single electron counting mode. A total of 17,010 movies were collected with a pixel size of 1.38 Å and total electron exposure of 51 e^-^/Å^2^ spread over 40 frames. Both datasets were acquired with a defocus range of −1.6 to −4 μm (Table S2).

### Cryo-EM image processing and real-space refinement

Micrograph movies were aligned with dose weighting applied using MotionCor2 (*40*), and the contrast transfer function (CTF) parameters were estimated from the frame sums with Gctf (*41*). Images exhibiting ice contamination or poor CTF estimation were discarded at this stage, leaving 15,803 (biliverdin complex) and 16,619 (Fab complex) movies for further processing. An initial set of particles, autopicked with Laplacian-of-Gaussian function in Relion-3.1 (*42*), was used to generate 2D class averages that served as templates for picking both datasets with Gautomatch (http://www.mrc-lmb.cam.ac.uk/kzhang/). Particles (1,209,334 and 2,505,265 for spike-biliverdin and spike-Fab, respectively) were extracted from weighted frame sums, binned 4-fold, and subjected to reference-free 2D classification in cryoSPARC-2 (*43*). 371,422 spike-biliverdin and 709,127 spike-Fab particles belonging to well-defined classes containing trimeric spike were selected for further processing (figs S4A, S11A). An initial 3D model was generated using *ab initio* procedure in cryoSPARC-2. Selected particles, re-extracted with 2-fold binning, were subjected to 3D classification into 11 (spike-biliverdin) and 16 (spike-Fab) classes in Relion-3.1. Particles belonging to selected 3D classes were re-extracted without binning and used in 3D reconstruction followed by CTF refinement (beam tilt and per-particle defocus) and Bayesian polishing in Relion-3.1. The final 3D reconstructions were done using non-uniform refinement procedure in cryoSPARC-2. Resolution is reported according to the gold-standard Fourier shell correlation (FSC), using the 0.143 criterion (*44, 45*) (figs S4C-D and S11C-D, Table S2). To aid in model building process, the maps were filtered and sharpened using deepEMhancer (*46*) or using density modification procedure in Phenix (*47*). An additional spike-Fab reconstruction was obtained using multibody refinement in Relion-3.1 accounting for two rigid bodies (one spanning the Fab moiety plus the associated NTD, and the second encompassing the rest of the structure) was used for illustration purposes (Fig. 4A).

For the 3RBD-down spike model, coordinates from protein databank entry 6ZGE (*13*) were docked into the cryo-EM map in Chimera (*48*). Residues 14-319 were replaced with the NTD crystal structure for each of the chains along with the associated biliverdin molecules. The model was manually adjusted in Coot (*49*) and refined using phenix.real_space_refine (version 1.19rc5-4047)(*50*). The model was then docked into the cryo-EM map of asymmetric 1RBD-up reconstruction and one RBD refitted to the extended position in Chimera; the model was manually adjusted in Coot and refined using phenix.real_space_refine. For the spike-Fab model, 3RBD-down spike model was fitted to the map using Chimera; the RBD of chain A was fitted in to the extended position, which is less well defined, and the NTD of chain C was extensively remodelled. The protein data bank was searched for similar structures using the sequence of the Fab chains; 6APC (*51*) and 6PHB (*52*) were selected as templates for the heavy and light chains of the Fab respectively, variable and constant sub-domains were fitted individually using either phenix.dock_in_map (*53*) and/or fitted in Chimera. The Fab fragments were manually adjusted to match the sequence of P0008_056 antibody and fitted to the cryo-EM map. The constant domains of the Fab are less well resolved in the map and minimal adjustments were made to these domains. The whole structure was subjected to automatic flexible fitting using Namdinator v2.13 (*54*) and then phenix.real_space_refine before another round of manual rebuilding in Coot and a final round of phenix.real_space_refine. Biliverdin (BLA) ligand geometry definition file was generated by Grade (Global Phasing) and model quality was assessed using Molprobity (*55*).

### Crystal Structure of the NTD in complex with biliverdin

Protein construct (spanning SARS-CoV-2 S1 residues 1-310) at 10 mg/ml was supplemented with 90 μM biliverdin before mixing with crystallization mother liquor in a 1:1 ratio. Plate-like crystals grew to 80-120 μm in two dimensions and ∼10-20 μm in the third dimension in conditions containing 24% PEG 3350 (w/v) and 0.25 M NaSCN by hanging drop vapour diffusion over 1-2 weeks at 18°C. Crystals were cryoprotected by the addition of PEG 400 to a final concentration of 30% (v/v) to the drop solution before flash freezing in liquid nitrogen.

X-ray diffraction data were collected at the PX1 beamline, Swiss Light Source, using wavelength 1 Å, 100% transmission, a 40-μm beam, 0.1-sec exposure and 0.5° rotation per image. Data were indexed, scaled and merged using XDS (*56*) and Aimless (*57*) via Xia2 (*58*). SARS-CoV-2 spike NTD (residues 14-290; PDB ID 6ZGE) (*13*) was used as a model for molecular replacement and yielded a solution containing one NTD per asymmetric unit, with a log likelihood gain of 490 and translation function Z-score of 22.7, in space group C222_1_ using Phaser (*59*) within the Phenix package (*53*). The initial molecular replacement solution was subjected to morph model in Phenix before commencing with rounds of manual fitting in Coot (*49*) and refinement using phenix.refine (version 1.19rc4-4035) (*53*). First, the protein chain was fitted and extended where possible, and refined, then glycosylation moieties were added where visualized in the positive *Fo-Fc* density, followed by conceivable PEG and water molecules. The electron density around the disulphide bonds suggested that they were labile and as such were modeled as alternative conformations between oxidized and reduced where appropriate and the occupancy refined between these states. The stability of the disulphide bonds could have been affected by trace amounts of DTT introduced during the treatment of the protein with 3C protease and EndoH. The R_free_ and R_work_ were 21.5 and 18.5%, respectively, before a biliverdin molecule was fitted into the prominent positive difference density. The final refinement included four Translation/Libration/Screw groups (residues 14-67, 68-202, 203-278, 279-319) that had been segmented by the TLSMD server (*60*). All ligand geometry definition files were generated by Grade (Global Phasing) and model quality was assessed using Molprobity (*61*). The final model consists of spike residues 14-319, one biliverdin molecule, seven N-liked glycans (attached to asparagine residues at positions 17, 61, 122, 149, 165, 234, and 282), 10 PEG moieties, and 351 water molecules and has reasonable geometry and fit to the electron density (Table S3, fig. S5B).

### Human sera

Following written informed consent, serum samples from staff and patients of the Imperial College Healthcare NHS Trust (ICHNT) and the Wellington Hospital diagnosed with SARS-CoV-2 infection were donated to the Communicable Diseases Research Tissue Bank (CDRTB) of the Section of Virology, Department of Infectious Disease, Imperial College London. The use of these sera was approved by the CDRTB Steering Committee in accordance with the responsibility delegated by the National Research Ethics Service (South Central Ethics Committee Oxford – C, NRES references 15/SC/0089 and 20/SC/0226). The median time from onset of symptoms (or positive RT-PCR test in the case of asymptomatic infection) was 29 (0-94) days. Additionally, serum or plasma samples were obtained from University College London Hospitals (UCLH) COVID-19 patients testing positive for SARS-CoV-2 infection by RT-qPCR and sampled between March 2020 and April 2020 (*18*). The sera were collected a median of 21 (9-31) days post onset of symptoms. Patient sera were from residual samples prior to discarding, in accordance with Royal College Pathologists guidelines and the UCLH Clinical Governance for assay development and approved by HRA (IRAS reference 284088). All serum or plasma samples were heat-treated at 56°C for 30 min prior to testing by flow cytometry.

### IgG capture assay

WT (depleted of biliverdin by chromatography under acidic conditions) and mutant SARS-CoV-2 S1 proteins (4.1 mg/ml; 100 μl) were conjugated to horse radish peroxidase (HRP) using the Lynx rapid HRP conjugation kit (BioRad). Following quenching and dilution in conjugate stabiliser (Clintech, Guildford, UK; product code #MI20080), half of each conjugate was supplemented with 10 μM biliverdin. Nunc 96-well, U8 MaxiSorp plates (Fisher Scientific) were coated overnight at 4°C with AffiniPure rabbit anti-human IgG antibody (Stratech; product code #309-005-008) diluted to 5 μg/ml in coating buffer (Clintech; product code #643005). Following a 3-h incubation at 37°C, and a 1-h incubation at room temperature, the wells were washed with washing buffer (Clintech; product code #20024) and incubated for 4 h in blocking solution (Clintech; product code #MI20011). The wells were air-dried and stored desiccated at 4°C until use. For ELISA, 100-μl serum samples, each diluted 1:100 in diluent buffer (Clintech, product code #2040), were added to the coated wells and incubated stationary at 37°C for 1 h. To detect S1-specific IgGs, the wells were washed with washing buffer (Clintech) and aspirated to dryness, following which 100 μl of S1-HRP fusion conjugate diluted in conjugate diluent (Clintech; product code #100171) to a previously defined optimum concentration (1:1,500) were added and incubated for one hour at 37°C. The wells were then washed as before and developed for 30 min at 37°C using tetramethylbenzidine substrate (Clintech; product code #2030b), quenched by the addition of 50 μl stop solution (Clintech; product code #20031). The resulting optical densities (ODs) were acquired using a SpectraMax M2 reader (Molecular Devices).

The IgG capture ELISA data was modelled with a Bayesian linear model, using the gamma likelihood function: Gamma(μ, Scale). The linear model took the form of log(μ) = intercept[Sample] + offset[Protein], where the intercept[Sample] term allows varying intercepts across samples (to account for the repeated measurements of each serum sample across conditions, assumed to be distributed as Gamma(*μ*_intercept_, scale_intercept_)), and the offset[Protein] term accounts for variation attributable to different protein coatings. Pairwise contrasts were drawn from the posterior distribution to construct credible intervals for the difference in OD values between different protein coatings. Priors: offset[Protein]∼Normal(−3,0.2), scale∼Exponential(5), *μ*_intercept_ ∼Normal(1,0.2), scale_intercept_ ∼Exponential(5). Monte Carlo settings: 10,000 iterations, 4 chains, adapt_delta = 0.95, sampler = NUTS.

### Flow cytometry

Serum antibody binding to full-length SARS-CoV-2 S expressed on HEK293T cells was performed using a recently described method (*18*). Briefly, HEK293T cells were transfected with an expression vector (pcDNA3) carrying codon-optimized genes encoding either the wild-type SARS-CoV-2 S (UniProt ID: P0DTC2) or N121Q SARS-CoV-2 S, using GeneJuice (EMD Millipore). Two days after transfection, cells were trypsinized and transferred into V-bottom 96-well plates (20,000 cells/well). Cells were incubated with sera (diluted 1:50 in PBS) for 30 min, with or without addition of 10 µM biliverdin throughout the staining period. They were then washed with FACS buffer (PBS, 5% BSA, 0.05% sodium azide) and stained with FITC anti-IgG (clone HP6017, Biolegend), APC anti-IgM (clone MHM-88, Biolegend) and PE anti-IgA (clone IS11-8E10, Miltenyi Biotech) for 30 min (all antibodies diluted 1:200 in FACS buffer). Samples were run on a Ze5 analyzer (Bio-Rad) running Bio-Rad Everest software v2.4, and analyzed using FlowJo v10 (Tree Star Inc.) analysis software. To calculate the effect of biliverdin, the mean fluorescence intensity (MFI) of positively stained cells only was used in each condition. The MFI data was modelled with a Bayesian linear model, using the gamma likelihood function: Gamma(μ, Scale). The linear model took the form of log(μ) = intercept[Sample] + offset[Protein], where the intercept[Sample] term allows varying intercepts across samples (to account for the repeated measurements of each serum sample across conditions, assumed to be distributed as Normal(*μ*_intercept_,*σ*_intercept_)), and the offset[Protein] term accounts for variation attributable to different protein coatings. Pairwise contrasts were drawn from the posterior distribution to construct credible intervals for the difference in MFI values between samples ±biliverdin. Priors: offset[Protein]∼Normal(0,400), scale∼Exponential(0.1), *μ*_intercept_∼Normal(2500,200), *σ*_intercept_∼Exponential(0.01). Monte Carlo settings: 10,000 iterations, 4 chains, adapt_delta = 0.95, sampler = NUTS.

The same procedure was performed to assess monoclonal IgG binding to cell surface SARS-CoV-2 spike with the following alterations: Transfection was performed with PEI-Max (1 mg/ml, Polysciences), FACS wash buffer (FWB) containing PBS supplemented with 1% BSA. Monoclonal IgGs were serially diluted 10-fold from 50 µg/ml prior to mixing with transfected cells. Antibody binding was detected with APC anti-IgG (Biolegend) diluted 1:200 in FWB buffer. Samples were run on a NovoCyte 96-well plate flow cytometer and analysed using FlowJo v10 (Tree Star) analysis software. Three buffer only samples and secondary antibody alone conditions were used to define the spike-positive gate.

### Monoclonal human antibodies

The following IgGs COVA1-26, COVA1-23, COVA2-38, COVA2-17, COVA1-20, COVA2-26, COVA1-22, COVA3-07, COVA2-03, COVA1-18,COVA1-12, COVA1-16, COVA2-01, COVA2-02, COVA2-04, COVA2-07, COVA2-11, COVA2-15, COVA2-29, COVA2-39, COVA2-44, COVA2-46, COVA2-10, COVA2-25, COVA2-30 have been reported (*10*). Patients P003, P008 and P0054 were part of the COVID-IP study (*62*). Cloning and characterisation of human IgGs P008_056, P003_027, P008_039, P008_051, P008_052, P003_014, P008_057, P008_100, P008_081, P008_017, P008_087, P054_021, P008_007, P003_055, and P008_108 will be described elsewhere (Graham *et al*., in preparation).

### ELISA with monoclonal IgGs

The assays were performed in a similar manner to the previously described protocol for serum samples (*18, 63, 64*). Briefly, high-binding ELISA plates (Corning, product code 3690) were coated with 3 µg/ml (25 µl per well) SARS-CoV2 WT S1 antigen (purified with or without acid treatment) or N121Q S1 in PBS, either overnight at 4 °C or for 2 h at 37 °C. Wells were washed with PBS supplemented with 0.05% Tween-20 (PBS-T) and blocked with 100 µl 2% casein in PBS for 1 h at room temperature. The wells were emptied and 25 µl of 2% casein in PBS was added per well. This solution was supplemented with biliverdin at 10 µM where indicated. Serial dilutions of IgGs were prepared in separate 96-well plate (Greiner Bio-One) in 2% casein, and then 25 µl of each serial dilution added to the ELISA assay plates and incubated for 2 h at room temperature. Wells were washed with PBS-T. Secondary antibody was added and incubated for 1 h at room temperature. IgG binding was detected using goat-anti-human-Fc conjugated to alkaline phosphatase (1:1,000; Jackson, product code 109-055-098). Wells were washed with PBS-T, and alkaline phosphatase substrate (Sigma-Aldrich) was added and read at 405 nm. Area under the curve values were calculated using GraphPad Prism.

### Pseudotype infectivity assay

Simian immunodeficiency (SIV) particles were used to assess effects of H207A, R190K and N121Q mutations on the function of SARS-CoV-2 spike. HEK293T cells, seeded one day earlier in 10-cm dishes in complete Dulbecco’s modified Eagle’s medium (DMEM, supplemented with 10% fetal bovine serum, FBS) were co-transfected with 17 µg SIVMAC239-GFP, an *env*- and *nef*-defective provirus construct expressing a GFP reporter (*65*), and 4 µg pcDNA-SARS-CoV-2-del19, encoding SARS-CoV-2 spike with or without mutations in the biliverdin binding pocket. To improve pseudotyping efficiency, the constructs encoded a truncation of the spike C-terminal 19 amino acids (*66*). Viral pseudotypes were harvested 48 h post-transfection, clarified by low-speed centrifugation at 300*g* for 5 min and filtered through a 0.45-μm filter. The stocks of viruses pseudotyped with the spike variants were diluted to an equal reverse transcriptase activity (*67*). Six 5-fold serially diluted virus stocks were inoculated in quadruplicate in 96-well plates onto Huh-7/ACE-2 and Vero/ACE2, modified to overexpress the receptor ACE-2 from a transduced lentiviral vector carrying the puromycin resistance gene, seeded one day before infection in 96-well plates; 48 h post-infection, fluorescent cells were counted using the Ensight plate reader (Perkin Elmer). Infectivity was calculated from values falling into a linear dilution range by dividing the number of infected cells in a well for the amount of reverse transcriptase activity associated to the virus inoculum, expressed in mU.

### Pseudotype neutralization assay

HIV-1 particles pseudotyped with SARS-Cov-2 spike were produced in a T75 flask seeded the day before with 3 million HEK293T/17 cells (American Type Culture Collection; catalogue code CRL-11268) in 10 ml complete DMEM, supplemented with 10% FBS, 100 IU/ml penicillin and 100 μg/ml streptomycin. Cells were transfected using 60 μg of PEI-Max (Polysciences) with a mix of three plasmids: 9.1 μg HIV-1 luciferase reporter vector (*63*), 9.1 μg HIV p8.91 packaging construct (*68*) and 1.4 μg WT SARS-CoV-2 spike expression vector (*18*) or its N121Q mutant version. Supernatants containing pseudotyped virions were harvested 48 h post-transfection, filtered through a 0.45-μm filter and stored at −80°C. Neutralization assays were conducted by serial dilution of monoclonal IgGs at the indicated concentrations in DMEM (10% FBS and 1% penicillin–streptomycin) and incubated with pseudotyped virus for 1 h at 37°C in 96-well plates. HeLa cells stably expressing ACE-2 (provided by J.E. Voss, Scripps Institute)(*69*) were then added to the assay (10,000 cells per 100 µl per well). After 48-72 h luminescence was assessed as a proxy of infection by lysing cells with the Bright-Glo luciferase kit (Promega), using a Glomax plate reader (Promega). Measurements were performed in duplicate and used to calculate 50% inhibitory concentrations (IC_50_) in GraphPad Prism software.

### SARS-CoV2 neutralization assay

Vero E6 (Cercopithecus aethiops derived epithelial kidney cells, provided by Prof. Wendy Barclay, Imperial College London) cells were grown in DMEM, supplemented with GlutaMAX (Thermo Fisher Scientific), 10% FBS, and 20 µg/ml gentamicin, and incubated at 37°C in 5% CO_2_ atmosphere. SARS-CoV-2 Strain England 2 (England 02/2020/407073) was obtained from Public Health England. The virus was propagated by infecting Vero E6 cells in T75 flasks (60-70% confluent), at a multiplicity of infection 0.005 in 3 ml of DMEM, supplemented with GlutaMAX and 10% FBS. Cells were incubated for 1 h at 37°C before adding 15 ml of the same medium. Supernatant was harvested 72 h post-infection following visible cytopathic effect, and filtered through a 0.22-µm filter, aliquoted and stored at −80C. The infectious virus titre was determined by plaque assay in Vero E6 cells.

Neutralization assays were performed as previously described (*63*). Cells were seeded at a concentration of 20,000 cells/100 μl per well in 96-well plates and allowed to adhere overnight. Serial dilutions of monoclonal antibodies were prepared with DMEM media (supplemented with 2% FBS, 100 IU/ml penicillin and 100 μg/ml streptomycin, Thermo Fisher Scientific) and incubated with SARS-CoV-2 for 1 h at 37°C. Biliverdin was added to the virus at a final concentration of 25 µM before addition to the antibody. The media was removed from the pre-plated Vero-E6 cells and the serum-virus mixtures were added to the Vero E6 cells and incubated at 37°C for 24 h. These virus/serum mixture were aspirated and cells fixed with 150 µl of 4% formaldehyde at room temperature for 30 min and then topped up to 300 µl with PBS. The cells were washed once with PBS and permeabilised with 0.1% Triton-X100 in PBS at room temperature for 15 min. The cells were washed twice with PBS and blocked using 3% milk in PBS at room temperature for 15 min. The blocking solution was removed cells were incubated with 2 µg/ml SARS-CoV-2 nuclear protein-specific murinized-CR3009 antibody in PBS supplemented with 1% milk at room temperature for 45 min. The cells were washed twice with PBS and horse anti-mouse-IgG-conjugated to HRP was added (1:2,000 in 1% milk in PBS, Cell Signaling Technology; product code S7076) at room temperature for 45 min. The cells were washed twice with PBS, developed using 3,3’,5,5’-tetramethylbenzidine substrate for 30 min and quenched using 2 M sulfuric acid prior to reading at 450 nm. Measurements were performed in duplicate.

## FIGURE LEGENDS

**Movie S1. Conformational changes in the NTD associated with binding neutralising antibody P008_056**.

**Figure S1.**
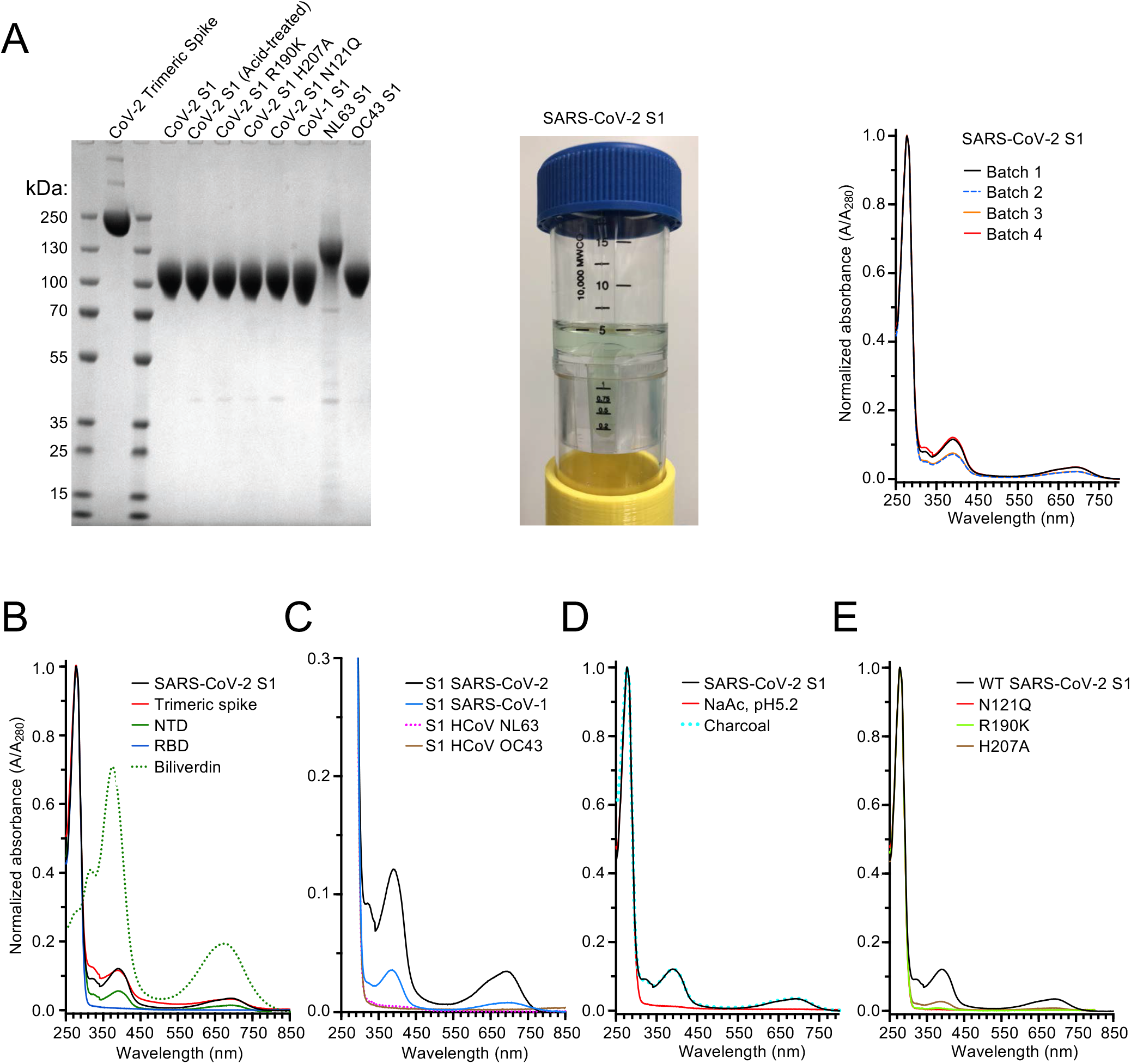
UV-visible light absorption spectral properties of coronaviral spike antigen constructs. (**A**) *Left:* Analysis of recombinant coronaviral spike and S1 constructs used in this work by SDS PAGE. Each lane contained 5 μg of purified recombinant protein; the gel was stained with Coomassie Blue. *Middle:* A representative SARS-CoV-2 S1 sample at ∼4 mg/ml in a centrifugal concentrator. *Right:* Light absorbance spectra of four independent SARS-CoV-2 S1 batches (250-800 nm). (**B-E**) Spectra of stabilised trimeric SARS-CoV-2 spike ectodomain (residues 1-1208), NTD (1-310), RBD (319-541), and biliverdin (**B**); SARS-CoV-1 S1 (residues 1-518), HCoV NL63 S1 (residues 1-664) and OC43 S1 (1-665) (**C**); SARS-CoV-2 S1 purified under acidic conditions in sodium acetate pH 5.2 or dialysed overnight against suspension of activated charcoal. Note the retention of biliverdin in dialysed sample, consistent with high affinity of the interaction at pH 8.0 (**D**); H207A, R190K and N121Q SARS-CoV-2 S1 (**E**), compared to a representative spectrum of WT SARS-CoV-2 S1 purified under standard conditions, shows as black lines. Spectra were acquired from proteins diluted to 1-1.2 mg/ml in HBSE buffer and normalised to absorption at 278 nm.

**Figure S2.**
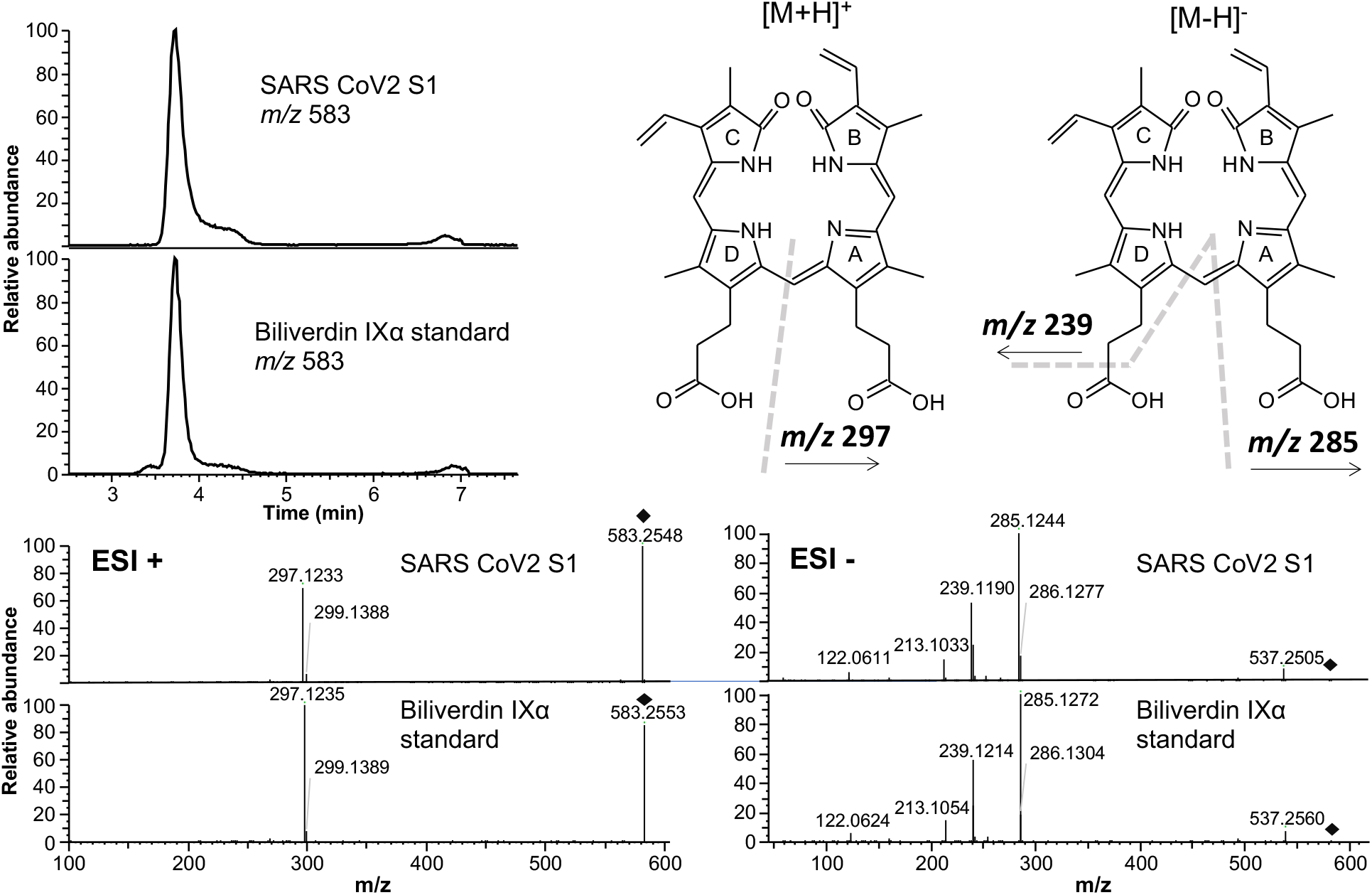
Identification biliverdin IXα by mass spectrometry. Positive-mode ion chromatograms of biliverdin XIα standard and of the pigment isolated from recombinant S1 (*top left*) and structure proposal for some of the fragment ions of biliverdin XIα (*top right*) The bottom panels show comparison of positive and negative MS2 spectra of the ions corresponding to biliverdin IXα in a standard (*upper*) and in recombinant S1 (*lower*). Black diamonds indicate expected positions of intact ions.

**Figure S3.**
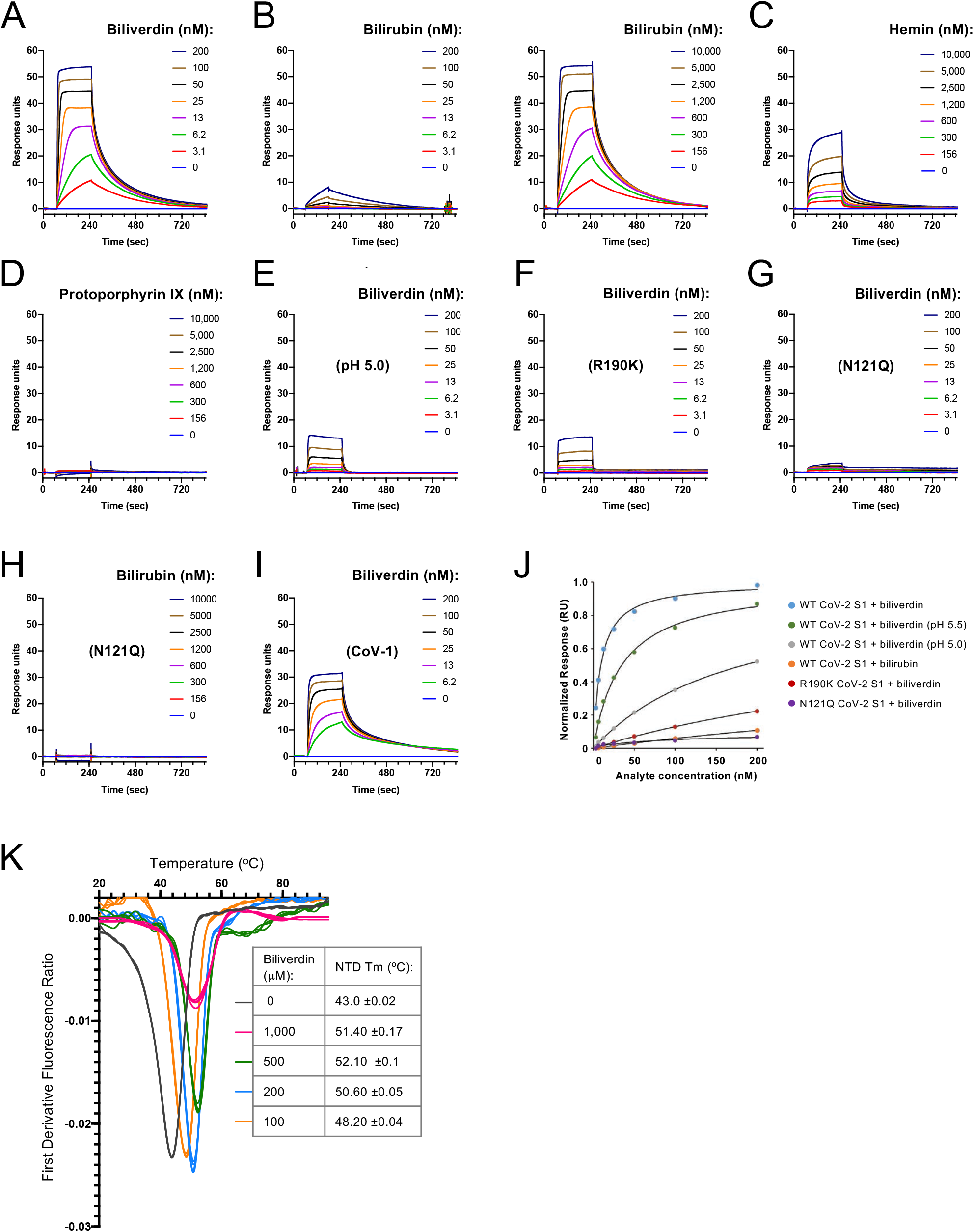
Representative SPR sensorgrams (A-I), estimates of equilibrium binding affinities (J) and melting point analysis (K). The sensorgrams were recorded with WT (**A-E**), R190K (**F**), or N121Q (**G-H**) SARS-CoV-2 S1, or WT SARS-CoV-1 S1 (**I**). The proteins were immobilised on a sensor chip and binding and dissociation of biliverdin IXα (**A, E-G, I**), bilirubin (**B, H**), hemin (**C**), and protoporphyrin IX (**D**) was measured. The analytes were injected at indicated concentrations at pH 8.0 (**A-D, F-I**) or pH 5.0 (**E**). The plots in panel **H** show representative normalised equilibrium binding curves; continuous black lines indicate fitting curves. Estimated K_d_s values are given in Table S1. Panel **K** shows melting behaviour of isolated SARS-CoV-2 NTD diluted to 30 μM in HBSE buffer (150 mM NaCl, 1 mM EDTA, 20 mM HEPES-NaOH, pH8.0) in the absence or presence of 100-1,000 μM biliverdin. The vertical axis corresponds to the first derivative of the ratios of fluorescence intensities measured at 350 and 330 nm wavelengths; the resulting melting points along with standard deviations (*n* = 4) are given in the inset.

**Figure S4.**
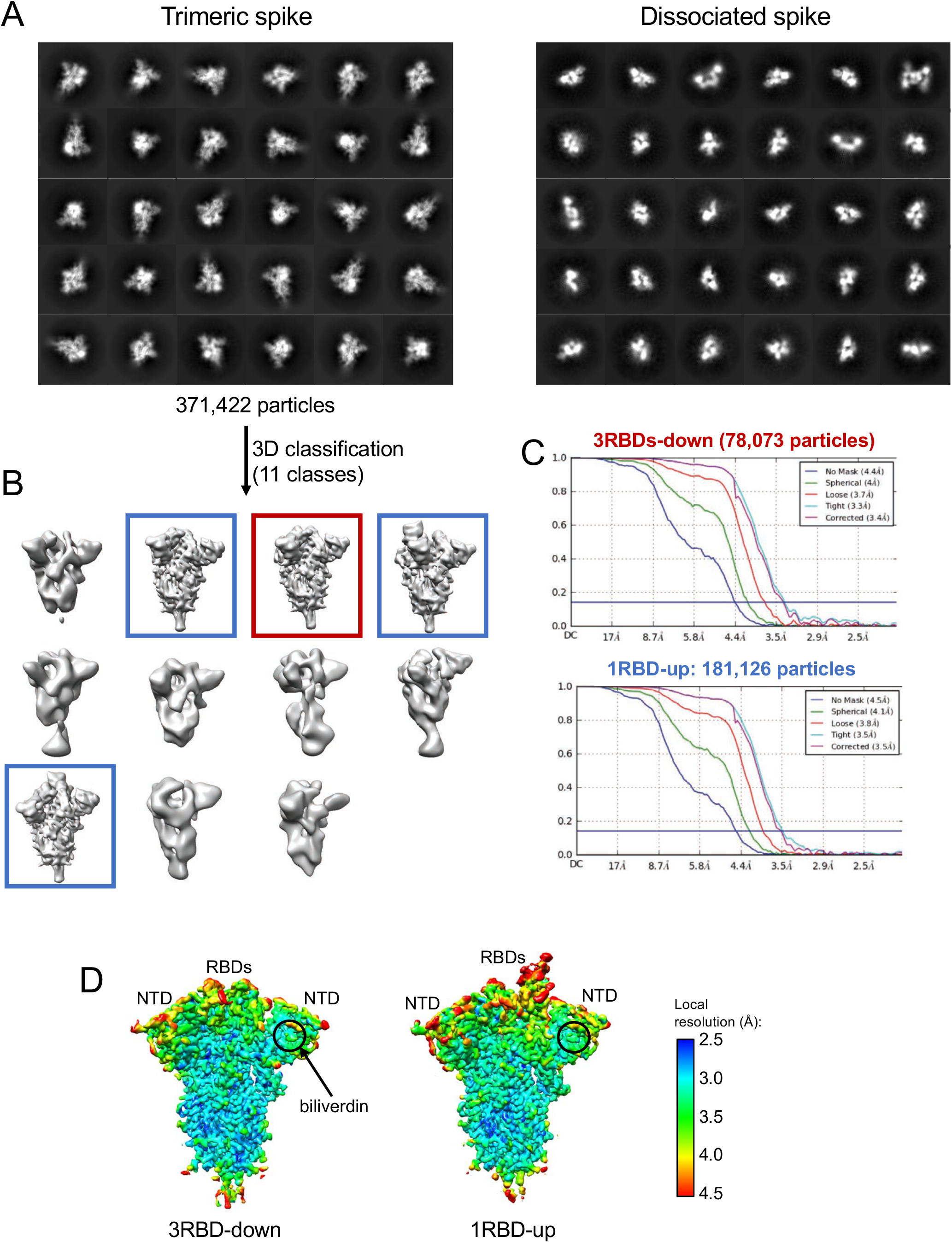
Cryo-EM image processing for the SARS-CoV-2 spike-biliverdin complex. (**A**) Representative 2D classes corresponding to trimeric (*left*) and dissociated (*right*) spike single particles. (**B**) Trimeric spikes identified by 2D classification (371,422 particles) were subjected to 3D classification in Relion-3.1 into 11 classes. Particles belonging to 3RBDs-down and 1RBD-up forms of the spike (indicated with red and blue boxes, respectively) were combined and used for 3D reconstruction in CryoSPARC-2. (**C**) Fourier shell correlations for the final reconstructions from CryoSPARC-2. (**D**) Maps colored by local resolution, as shown on the inset.

**Figure S5.**
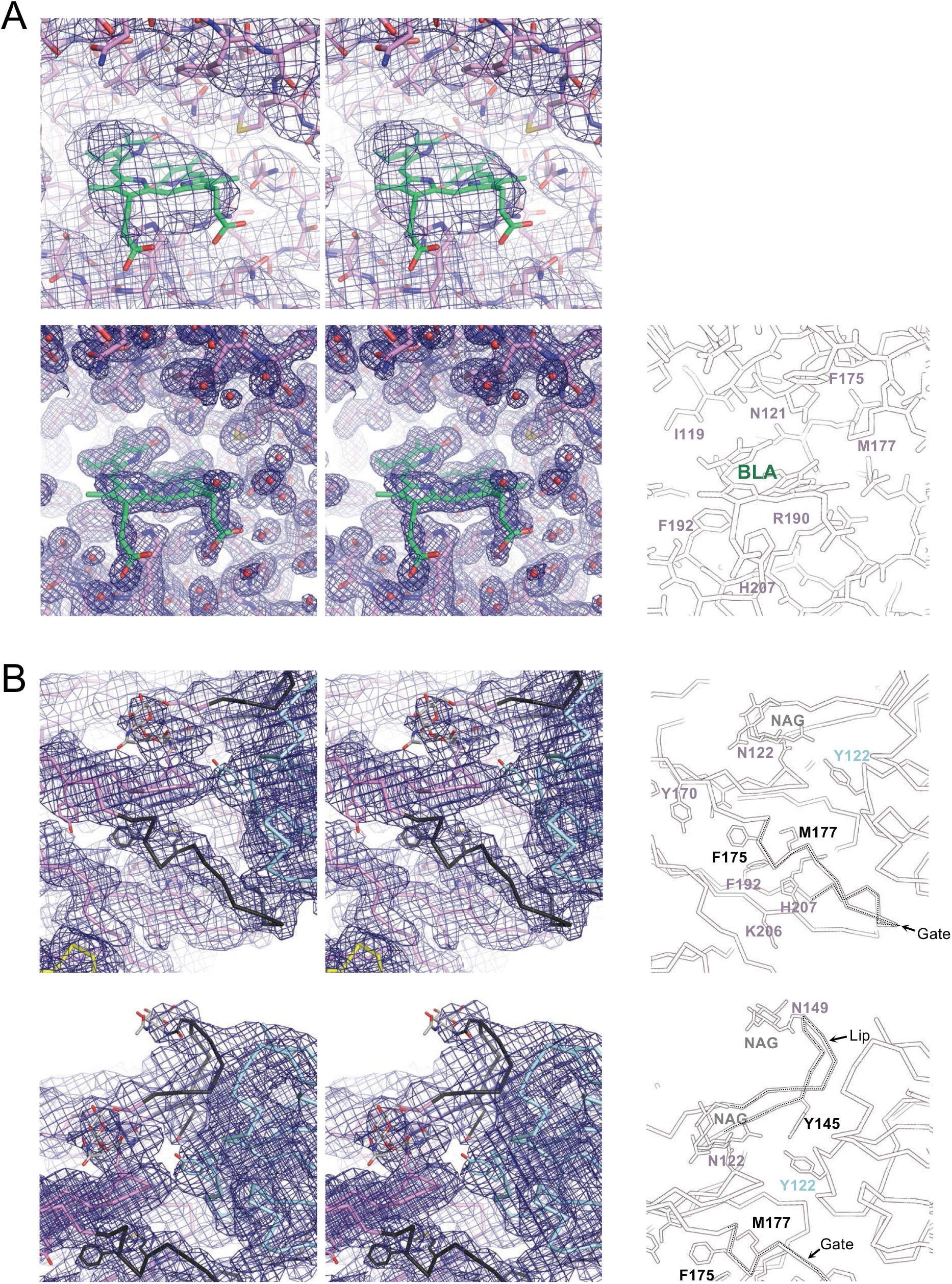
Stereo views of the cryo-EM and crystal structure density maps. Panel **A** shows the cryo-EM map of the 3RBD-down spike with biliverdin (*top*) and *2Fo-Fc* map of NTD-biliverdin complex crystal structure (*bottom*). The crystal structure is shown as sticks, coloured as in Fig. 1. Selected amino acid residues are indicated on the black-and-white image (*bottom right*). Electron density is contoured at r.m.s.d. of 1.0. Panel **B** shows cryo-EM map of the spike-Fab complex. Protein chains are depicted as ribbons, colored as in Fig. 4. Selected side chains are shown as sticks. Positions of selected residues, the gate and the lip are indicated on the back-and-white images to the right.

**Figure S6.**
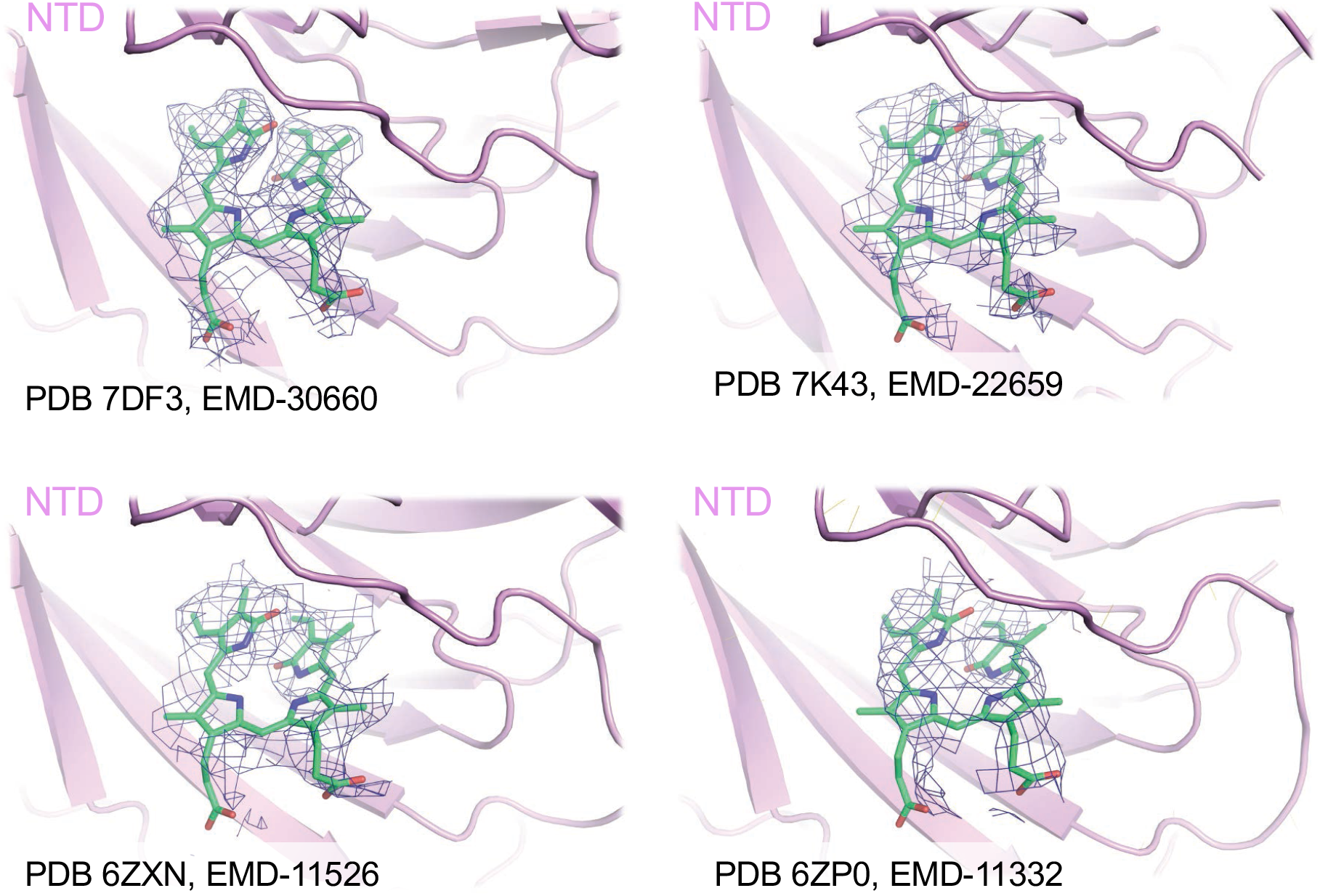
Evidence of biliverdin presence in previous SARS-CoV-2 spike cryo-EM reconstructions. Four published structures (*14-17*) with map features associated with the NTD and not explained by the original models are shown as chicken wire. In each case, a biliverdin molecule was placed by superposition of the NTD-biliverdin complex crystal structure.

**Figure S7.**
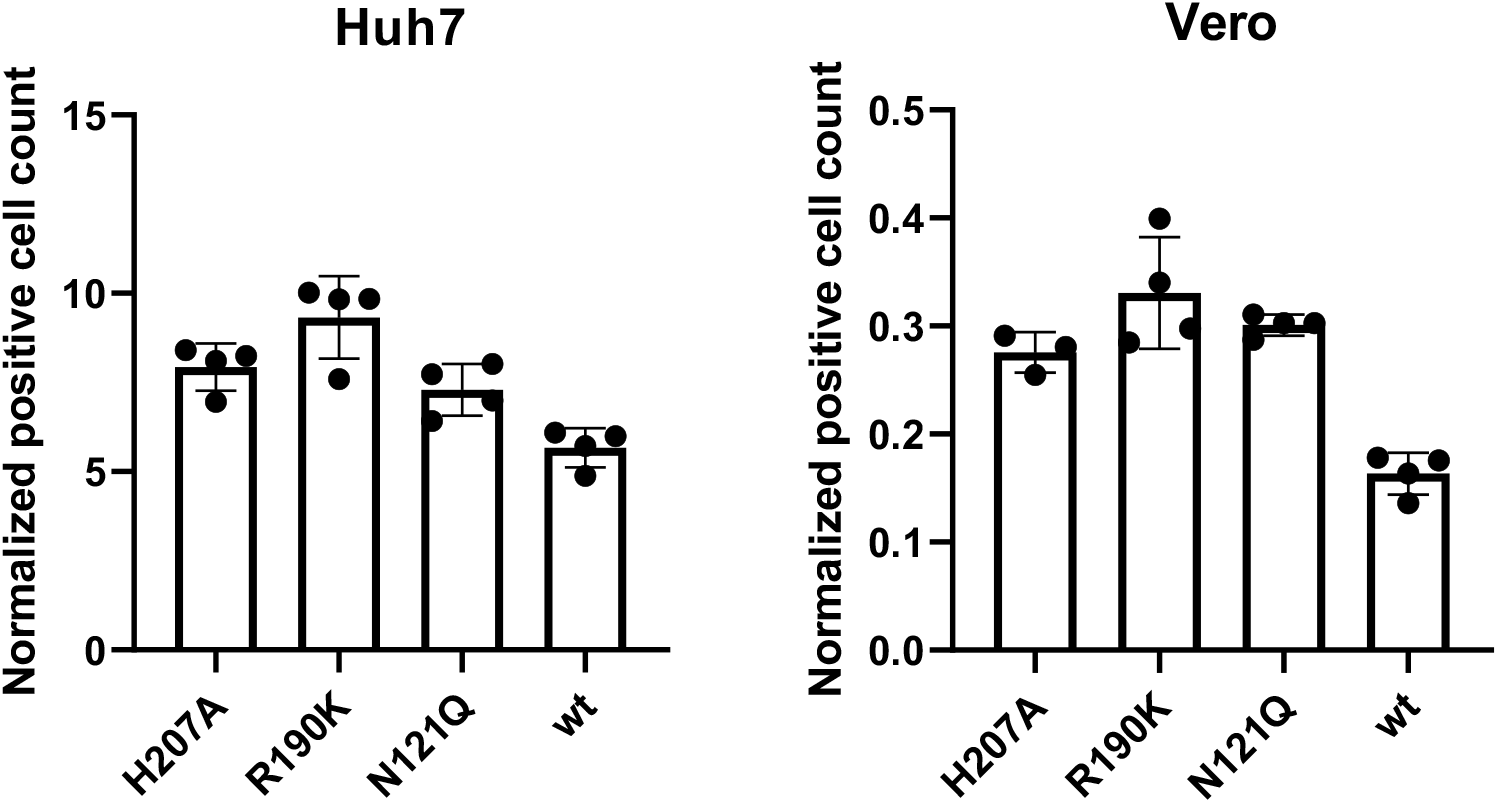
Infectivity of lentiviral vectors pseudotyped with SARS-CoV-2 spike with and without indicated mutations within the NTD on Vero (*left*) and Huh7 (*right*) cells. Infectivity is expressed as number of infected cells relative to the amount of RT-activity associated to the virus inoculum. Bars represent means and data from individual measurements (n = 4) are shown with dots. Error bars represent standard deviation of the mean calculated from the quadruplicate determinations.

**Figure S8.**
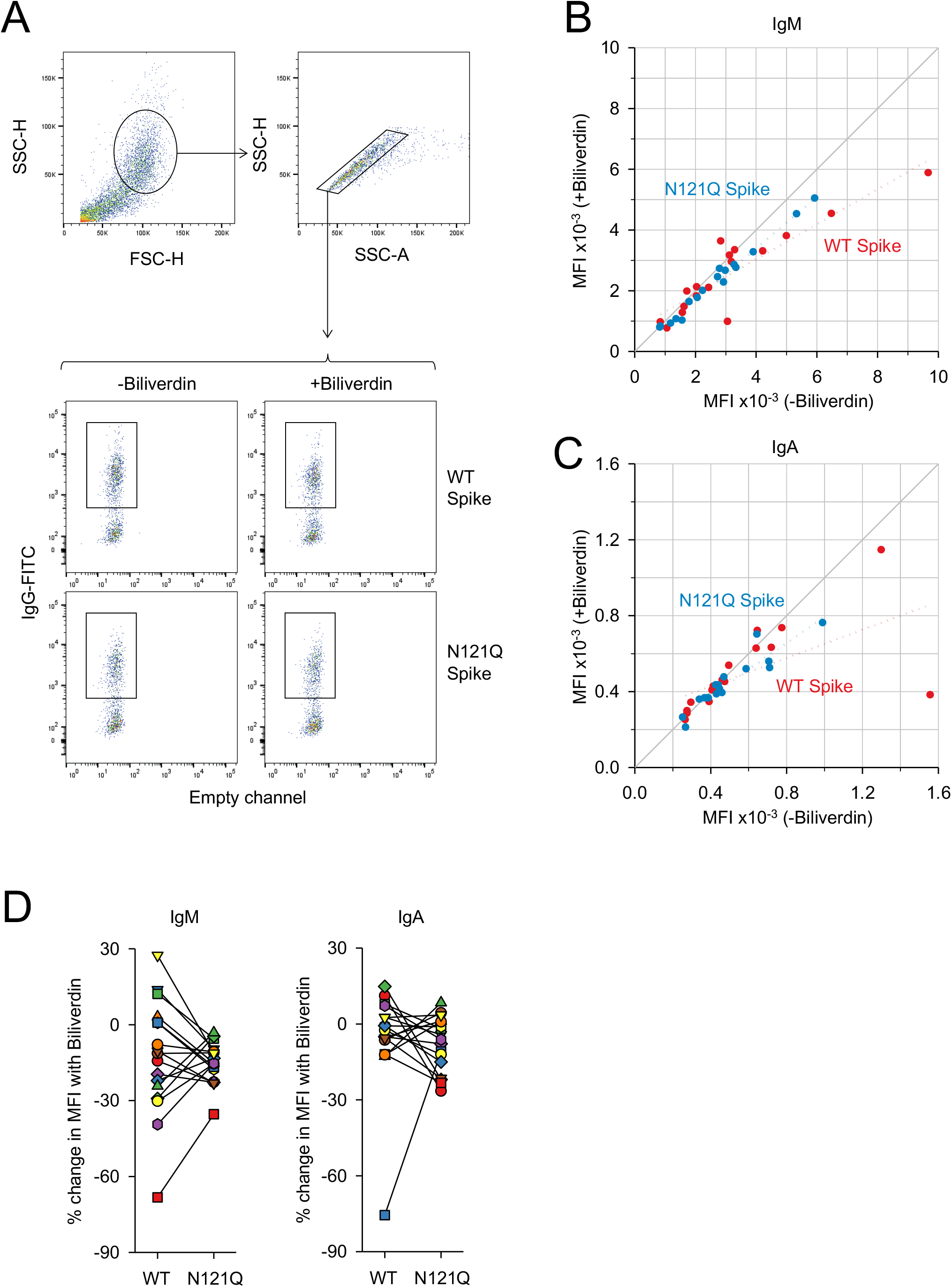
(**A**) Gating strategy of HEK293T cells transfected to express WT spike or N121Q SARS-CoV-2 spike. Staining intensity was taken as the MFI of positively stained cells, represented by the rectangular gates. (**B-C**) MFI of IgM (**B**) and IgA (**C**) staining of HEK293T cells expressing WT or N121Q SARS-CoV-2 spike by individual patient sera in the absence or the presence of 10 μM biliverdin. Each symbol represents an individual patient (n=17) and coloured dotted lines represent the linear regression for each spike variant. (**D-E**) Change in MFI caused by the addition of 10 μM biliverdin, as percent of staining without biliverdin, for serum for IgM and IgA antibodies. Each pair of connected symbols represents an individual patient.

**Figure S9.**
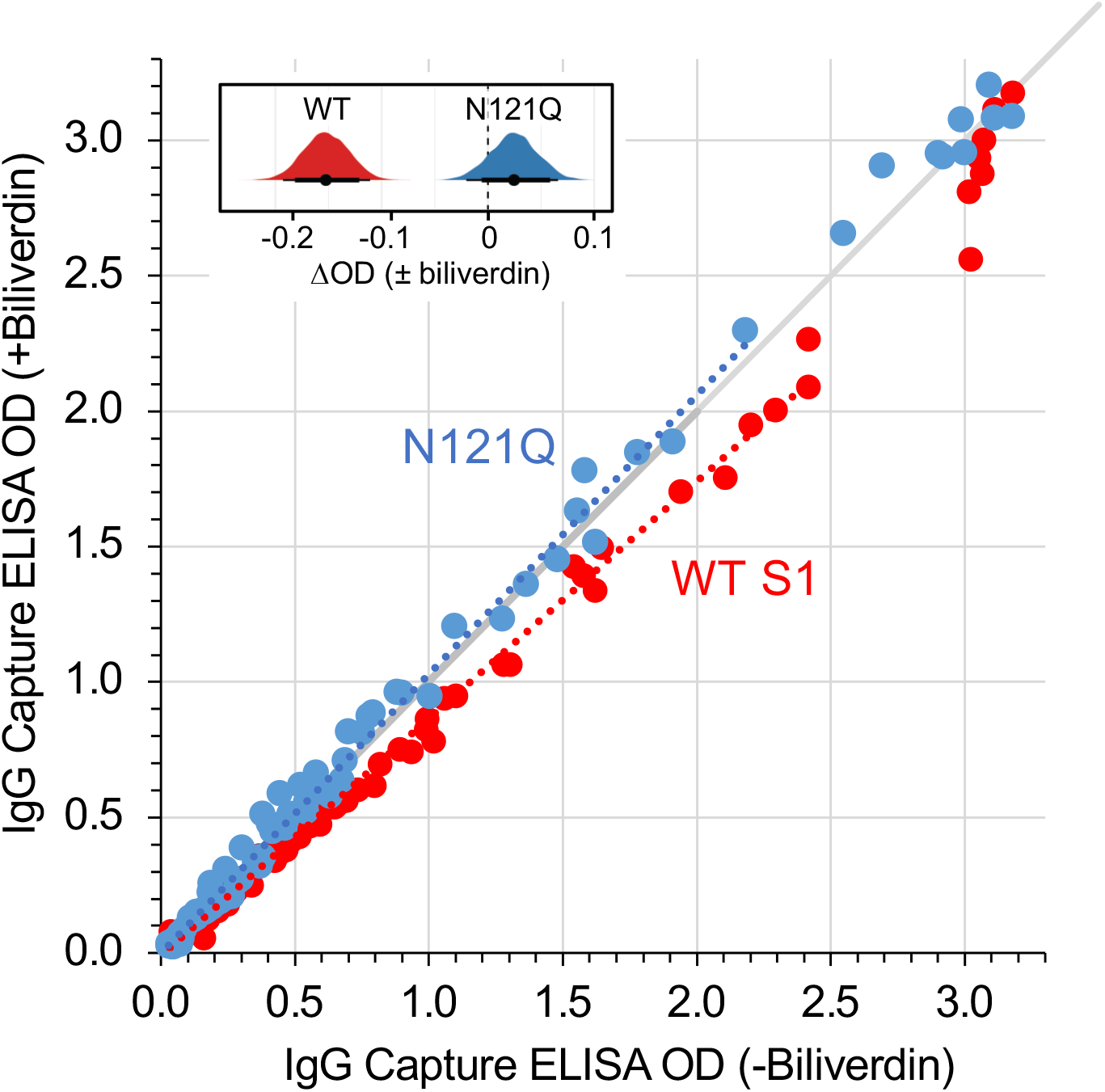
Effect of biliverdin binding on reactivity of SARS-CoV-2 with convalescent sera. IgG capture ELISA was performed with recombinant WT (biliverdin-depleted, red circles) or N121Q (blue circles) S1 antigen using sera from SARS-CoV-2 infected and convalescent individuals (n=91) in the presence (vertical axis) or absence (horizontal axis) of 10 μM biliverdin. Dotted lines represent linear fit for each set (considering samples with OD values <2.5, n = 82); the grey line is the diagonal. The inset shows posterior probability density plots of values for pairwise contrasts (±biliverdin) for the WT and N121Q S1 proteins. Black dots indicate the median of the distribution, thick and thin line ranges correspond to the 85% and 95% highest density interval, respectively; the dotted vertical line indicates a zero difference. Antibodies binding to S1 epitopes, which are occluded within protein-protein interfaces of the full-length trimeric spike, contribute to the observed signal in this ELISA assay. Therefore, the signal differences (±biliverdin) are expected to be less pronounced than in the flow cytometry assay, which used the complete native spike (Fig. 2).

**Figure S10.**
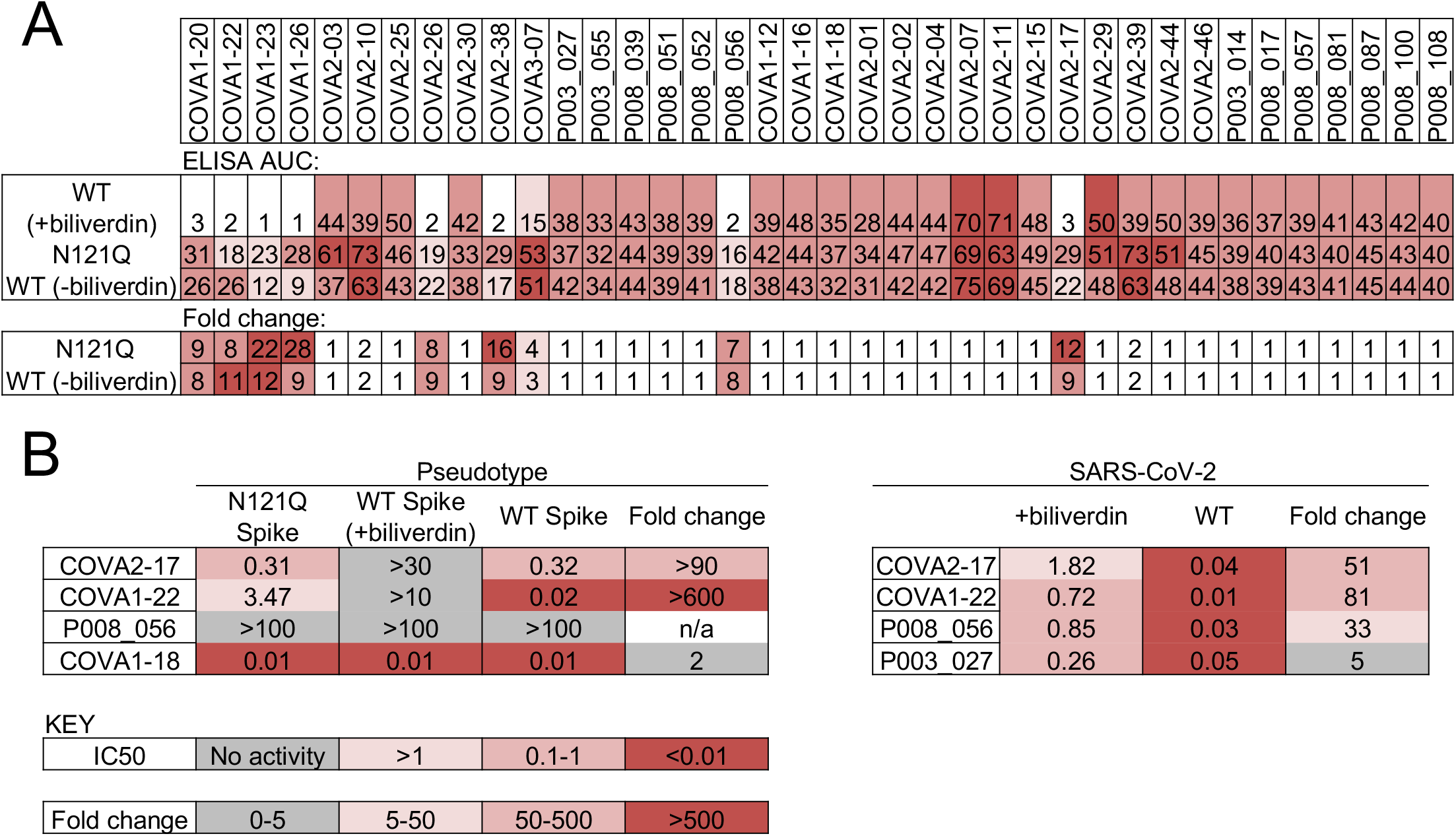
Biliverdin decreases binding to SARS CoV-2 spike by a group of human monoclonal IgGs. (**A**) Antibodies observed previously to bind to spike were titrated 6-fold and assayed by IgG ELISA for binding to WT S1 or N121Q S1. WT protein was prepared under acidic conditions to remove any co-purifying biliverdin, and was used in the absence (-biliverdin) or presence (+biliverdin) of 10 μM exogenous biliverdin. Area under the curve (AUC) is shown for 38 IgG that bound to S1 (also see Figs 3A and 3C). AUC values are colour-coded as per the key. Fold change compared to WT S1 protein are reported. (**B**) IC_50_ values in μg/ml calculated in Graphpad Prism are shown for each combination of IgG and viral preparation. The fold change column shows the decrease in IC_50_ with BLV relative to either wild-type S pseudotyped or live virus. Colour-coding is indicated in the key.

**Figure S11.**
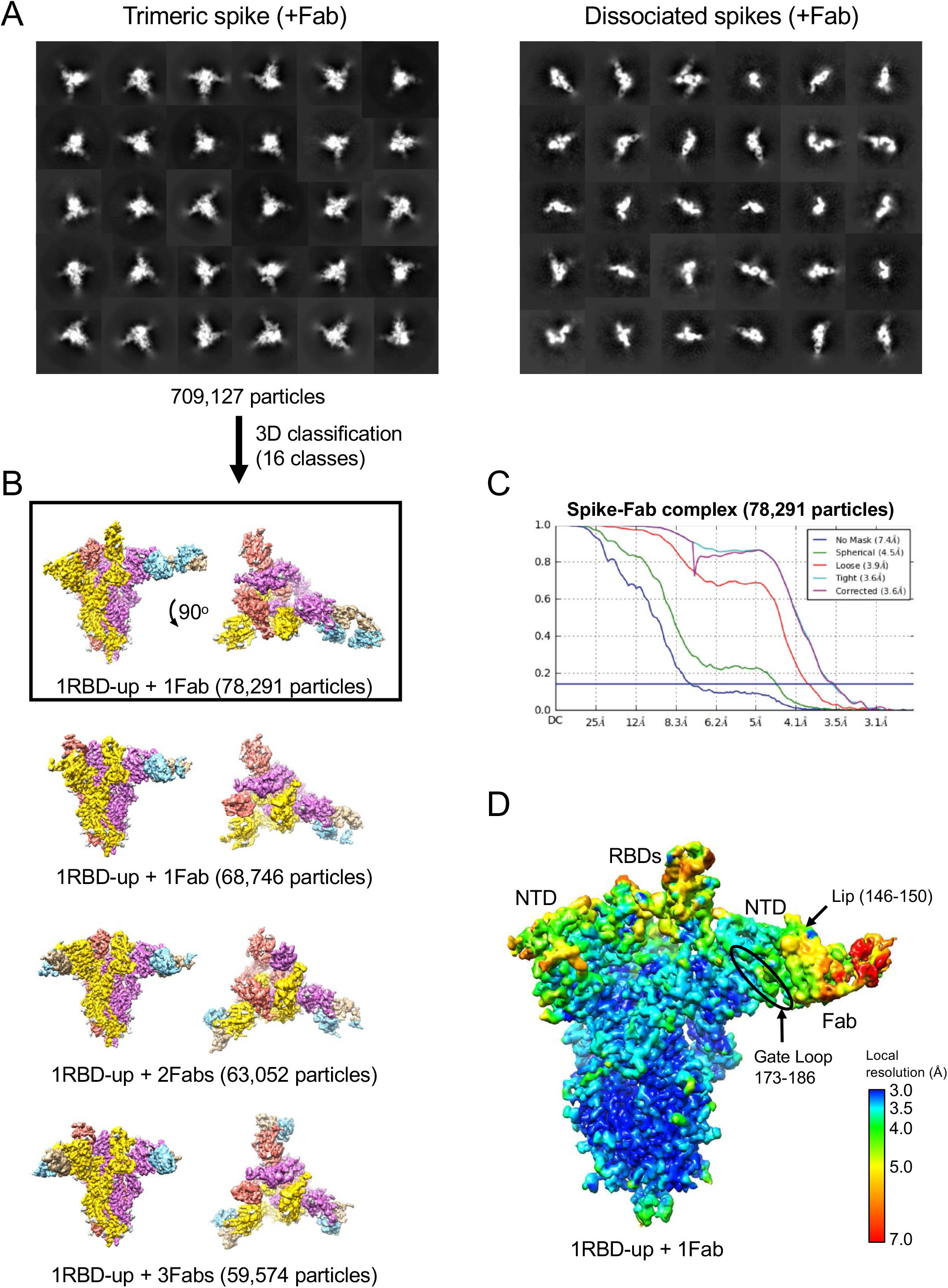
Cryo-EM image processing for the spike-Fab complex. (**A**) Representative 2D classes corresponding to trimeric (*left*) and dissociated (*right*) spike single particles. (**B**) Trimeric spikes identified by 2D classification (709,127 particles) were subjected to 3D classification in Relion-3.1 into 16 classes. Particles belonging to a class corresponding to trimeric spike (1RBD-up) with single Fab bound (boxed) were used in final 3D reconstruction in CryoSPARC-2. (**C**) Fourier shell correlations for the final reconstructions from CryoSPARC-2. (**D**) Cryo-EM map colored by local resolution.

**Table S1.** Affinity of SARS CoV2 S1 interaction with tetrapyrroles measured using surface plasmon resonance.

| S1 protein | Ligand | Buffer<sup>1</sup> | Kd (nM)<sup>2</sup> | Mean Kd (nM)<sup>3</sup> |
| --- | --- | --- | --- | --- |
| WT, CoV-2 | Biliverdin | Hepes pH 8.0 | 8.0; 9.2 | 9.8 ±1.3 |
| WT, CoV-2 | Biliverdin | 1% DMSO, Hepes pH 8.0 | 10.6; 11.5 |  |
| WT, CoV-2 | Biliverdin | 1% DMSO, Hepes pH 7.0 | 13.5; 14.5 |  |
| WT, CoV-2 | Biliverdin | 1% DMSO, BTP pH 6.0 | 13.6; 14.2 |  |
| WT, CoV-2 | Biliverdin | 1% DMSO, BTP pH 5.5 | 34.5; 29.9 |  |
| WT, CoV-2 | Biliverdin | 1% DMSO, BT pH 5.0 | 186.8; 194.1; 358.5 | 250 ±100 |
| WT, CoV-2 | Bilirubin | 1% DMSO, Hepes pH 8.0 | 540; 620; 1,000 | 720 ±250 |
| WT, CoV-2 | Hemin | 1% DMSO, Hepes pH 8.0 | 5,800; 6,640; 8,200 | 7,000 ±1,200 |
| WT, CoV-2 | Protoporphyrin | 1% DMSO, Hepes pH 8.0 | >10,000 | ` |
| WT, CoV-1 | Biliverdin | Hepes pH 8.0 | 18.7; 19.6; 20.4 | 19.6 ±0.8 |
| R190K, CoV-2 | Biliverdin | Hepes pH 8.0 | 1,500; 2,230 |  |
| N121Q, CoV-2 | Biliverdin | Hepes pH 8.0 | 16,800; 19,200 |  |
| N121Q, CoV-2 | Bilirubin | 1% DMSO, Hepes pH 8.0 | >10,000 |  |

<sup>1</sup>Running buffer variables (DMSO and pH) are shown; full composition is given in Methods section. BTP, BisTris Propane; BT, BisTris.
<sup>2</sup>Results of individual experiments performed in equilibrium mode.
<sup>3</sup>Mean and standard deviation shown for $N > 2$ measurements.

**Table S2.** Cryo-EM data collection, refinement and validation statistics

|  | SARS CoV-2 Spike<br>+ biliverdin |  | SARS CoV-2 Spike<br>+ P008_056 Fab |
| --- | --- | --- | --- |
| **Data collection** |  |  |  |
| Magnification | 128,440 |  | 36,232 |
| Voltage (keV) | 300 |  | 300 |
| Electron exposure (e<sup>-</sup>/Å<sup>2</sup>) | 33.6 |  | 51 |
| Defocus range (μm) | -1.6 to -4 |  | -1.6 to -4 |
| Pixel size (Å) | 1.09 |  | 1.38 |
| **3D reconstruction** |  |  |  |
| Molecular assembly | 3RBDs-down | 1RBD-up | 1RBD-up + 1Fab |
| Initial particle images (no.) | 1,209,334 | 1,209,334 | 2,505,265 |
| Final particle images (no.) | 78,073 | 181,126 | 78,291 |
| Symmetry imposed | C3 | C1 | C1 |
| Map resolution (Å) | 3.36 | 3.5 | 3.6 |
| FSC threshold | 0.143 | 0.143 | 0.143 |
| Map resolution range (Å) |  |  |  |
| **Refinement** |  |  |  |
| Model resolution (Å) | 3.36 | 3.5 | 3.6 |
| Model resolution range (Å) | 2.5-4.5 | 2.5-6.0 | 3.0-7.0 |
| Map sharpening *B* factor (Å<sup>2</sup>) | -161 | -156 | -66 |
| Model composition |  |  |  |
| Nonhydrogen atoms | 26,959 | 26,805 | 30,006 |
| Protein residues | 3,261 | 3,261 | 3,698 |
| Ligands | BLA: 3;<br>NAG: 84 | BLA: 3,<br>NAG:77 | BLA: 2,<br>NAG:79 |
| *B* factors (Å<sup>2</sup>) |  |  |  |
| Protein | 56.84 | 58.43 | 158.38 |
| Ligand | 94.84 | 91.87 | 147.91 |
| R.m.s. deviations |  |  |  |
| Bond lengths (Å) | 0.003 | 0.003 | 0.004 |
| Bond angles (°) | 0.521 | 0.587 | 0.552 |
| CC | 0.794 | 0.793 | 0.82 |
| **Validation** |  |  |  |
| MolProbity score | 1.59 | 1.66 | 1.84 |
| Clashscore | 6.08 | 6.86 | 7.28 |
| Poor rotamers (%) | 0.00 | 0.00 | 0.03 |
| Ramachandran plot |  |  |  |
| Favored (%) | 96.23 | 95.8 | 93.11 |
| Disallowed (%) | 0 | 0 | 0.16 |

**Table S3.** X-ray data collection and refinement statistics.

|  | NTD with biliverdin |
| --- | --- |
| **Data collection:** |  |
| Wavelength (Å) | 1.00 |
| Space group | C222<sub>1</sub> |
| **Unit cell parameters** |  |
| *a*, *b*, *c* (Å) | 98.07, 100.19, 85.97 |
| $\alpha$, $\beta$, $\gamma$ (°) | 90, 90, 90 |
| Number of crystals used | 1 |
| Resolution (Å) | 50.10-1.82 (1.87-1.82) <sup>a</sup> |
| **Number of reflections** |  |
| measured | 510,757 (34,535) |
| unique | 38,068 (2,758) |
| Completeness (%) | 99.4 (98.6) |
| Multiplicity | 13.4 (12.5) |
| $\langle I/\sigma(I) \rangle$ | 12.4 (1.1) |
| *R*<sub>merge</sub> (%) | 0.166 (2.407) |
| *R*<sub>pim</sub> (%) | 0.048 (0.721) |
| CC 1/2 | 0.999 (0.408) |
| **Refinement statistics:** |  |
| Resolution range (Å) | 49.03-1.82 (1.885-1.82) |
| **Number of reflections** |  |
| total | 38,038 (3,741) |
| free | 1,917 (165) |
| *R*<sub>work</sub>/*R*<sub>free</sub> | 0.1636/0.1997 |
| **Number of atoms** |  |
| total | 3,096 |
| protein | 2,511 |
| ligands | 234 |
| solvent | 351 |
| **R.m.s. deviations from ideal** |  |
| bond lengths (Å) | 0.01 |
| bond angles (°) | 1.0 |
| Average B-factor (Å<sup>2</sup>) | 32.76 |
| Clash Score <sup>b</sup> | 5.32 |
| Favored Rotamers <sup>b</sup> | 95.1 |
| **Ramachandran plot (%) <sup>b</sup>** |  |
| favored | 96.05 |
| disallowed | 0.00 |

<sup>a</sup> Values in parentheses correspond to the highest resolution bin.
<sup>b</sup> Analyzed using MolProbity (http://molprobity.biochem.duke.edu/).

## Notes

### Competing Interest Statement

The authors have declared no competing interest.

### Funding Statement

This research was funded by the Francis Crick Institute (FC001061, PC), which receives its core funding from Cancer Research UK, the UK Medical Research Council, and the Wellcome Trust. It was funded also by US National Institutes of Health (grant number AI150481, PC) the Kings Together Rapid COVID-19 Call award (KJD and MHM), the Huo Family Foundation (MHM and KJD) and by the UCL Coronavirus Response Fund made possible through generous donations from UCL supporters, alumni and friends (LEM, LM). Flow cytometry was supported by a multi-user equipment grant from The Wellcome Trust to MHM and KJD (208354/Z/17/Z). LEM is supported by a Medical Research Council Career Development Award (MR/R008698/1). MJvG is a recipient of an AMC Fellowship, and RWS is a recipient of a Vici grant from the Netherlands Organization for Scientific Research (NWO). CG was supported by the MRC-KCL Doctoral Training Partnership in Biomedical Sciences (MR/N013700/1). MOM, EP and RST were supported in part by an UKRI/MRC Covid-19 grant (MC_PC_19078). EN, MJS, JH receive support from the UKRI COVID-19 research scheme. This study is part of the EDCTP2 program supported by the European Union (grant number RIA2020EF-3008 COVAB) (KJD and MHM). The views and opinions of authors expressed herein do not necessarily state or reflect those of EDCTP. This research was funded in whole, or in part, by the Wellcome Trust. For the purpose of Open Access, the author has applied a CC BY public copyright license to any author accepted manuscript version arising from this submission.

### Author Declarations

The use of these sera was approved by the CDRTB Steering Committee in accordance with the responsibility delegated by the National Research Ethics Service (South Central Ethics Committee Oxford C, NRES references 15/SC/0089 and 20/SC/0226). Additionally, serum or plasma samples were obtained from University College London Hospitals (UCLH) COVID-19 patients testing positive for SARS-CoV-2 infection by RT-PCR and sampled between March 2020 and April 2020. Patient sera were from residual samples prior to discarding, in accordance with Royal College Pathologists guidelines and the UCLH Clinical Governance for assay development and approved by the Health Research Authority (HRA, IRAS reference 284088).

